# The Impact of SARS-CoV-2 Vaccine Dose Separation and Dose Targeting on Hospital Admissions and Deaths from COVID-19 in England

**DOI:** 10.1101/2022.08.22.22278973

**Authors:** Matt J. Keeling, Samuel Moore, Bridget Penman, Edward M. Hill

## Abstract

In late 2020, the JCVI (the Joint Committee on Vaccination and Immunisation, which provides advice to the Department of Health and Social Care, England) made two important recommendations for the initial roll-out of the COVID-19 vaccine. The first was that vaccines should be targeted to the elderly and vulnerable, with the aim of maximally preventing disease rather than infection. The second was to increase the interval between first and second doses from 3 to 12 weeks. Here, we re-examine these recommendations through a mathematical model of SARS-CoV-2 infection in England. We show that targeting the most vulnerable had the biggest immediate impact (compared to targeting younger individuals who may be more responsible for transmission). The 12-week delay was also highly beneficial, estimated to have averted between 32-72 thousand hospital admissions and 4-9 thousand deaths over the first ten months of the campaign (December 2020 - September 2021) depending on the assumed interaction between dose interval and efficacy.

## Introduction

Without doubt, the development of vaccines has been one of the greatest breakthroughs in the fight to reduce the scale and impact of the on-going SARS-CoV-2 pandemic. Following the deployment of vaccination programmes in late 2020 and early 2021 many countries were able to relax mitigation measures. Vaccines need to be deployed effectively to maximise their public health benefit; this requires detailed epidemiological input from a variety of sources, often extrapolated through mathematical projections. However, decisions need to be made rapidly to have the maximal benefit, usually before all the evidence can be systematically gathered and collated.

The UK was the first country to begin their vaccine deployment campaign, rapidly followed by Israel, the U.S.A. and European nations [1]. One of the earliest decisions was to target vaccination at the most vulnerable groups (to generate the maximal impact on severe disease) and healthcare workers (to maintain a functioning healthcare system); these prioritisation decisions for the initial roll-out of the COVID-19 vaccine in England followed recommendations from the JCVI (the Joint Committee on Vaccination and Immunisation) [2]. An alternative prioritisation programme may instead have sought to reduce infection as quickly as possible [3], by vaccinating the younger generations that were the most notable group in driving transmission. The general question as to when vaccination should be targeted at individuals generating the most transmission and when it should be targeted at the vulnerable remains an open problem (and is likely to depend on population demographics, vaccine characteristics and the heterogeneity in transmission risk and disease severity); but in late 2020 with an absence of data about the extent that the vaccine would be effective in reducing transmission, the decision to vaccinate the elderly and vulnerable first was relatively clear cut [4]. Most other countries adopted a similar targeted approach, although there were subtle differences in the prioritising of particular vulnerabilities, age-groups and at-risk professions. For example, the UK’s first four priority groups were: care home residents and workers; those over 80 and health/social care workers; those over 75; those over 70 or clinically extremely vulnerable [2] - which was in very close agreement with the first two priority groups in Germany [5]. The French priority order was similarly age-structured (care home residents and care home workers over 50; care-givers over 50 and firefighters; those over 75 years old; those age 65-74; those age 50-64 together with those with co-morbidities, essential sector workers and those with contact with the public [6]) as was the order in the USA (health care workers and longterm care facility residents; persons aged over 75 years and front-line essential workers; persons aged 65–74 years, persons aged 16–64 years with high-risk medical conditions, and any essential workers not included in the initial phases [7]).

Related to the idea of offering maximal early protection to the elderly and vulnerable was the question of whether to prioritise first or second doses. At the time, both vaccine supply and deployment capacity were limited, and the UK was experiencing another surge in cases due to the Alpha variant [8], so making maximal use of available resources was key. Essentially the question can be conceptualised as whether it is better to provide maximal two-dose protection to a smaller number of the most vulnerable first, or whether to prioritise lower one-dose protection to a greater number of individuals [9]. For example, using population sizes from England, is it better to give two doses and hence 80% protection to the ∼ 500,000 people over 90 years of age or one dose and therefore 50% protection to the ∼ 1,000,000 people over 87; for real-world decisions this caricature is made more complex by different degrees of vulnerability and changing vaccination capacity (which was generally increasing throughout the early phase following initial roll-out). In addition, evidence was emerging that the AstraZeneca ChAdOx1 vaccine, which was the main early component of the UK programme, had better efficacy if the interval between doses was extended from 3 to 12 weeks [10]. The decision was therefore made in the UK to adopt a 12-week dose interval for all UK vaccines [11], allowing the first three months of the programme to be dedicated to giving as many people as possible the protection offered by one dose of the vaccine. Very few countries followed this example; most including the USA and the majority of countries in Europe opted for a 3-4 week interval [12, 13] although Canada instigated a 16 week interval [14]. (We note that in May 2021, to combat the rise in hospital admissions, the dose interval in the UK was shortened to 8-weeks [15], in response to concerns about the degree of protection from one dose against the Delta variant and to ensure two-dose protection in more vulnerable groups.)

## Results

Here we use mathematical models, developed and matched to epidemic data from England throughout the pandemic (see Supplementary Material), to retrospectively assess the implications of the vaccine dose separation and prioritisation decisions by simulating counterfactual scenarios. We consider four different scenarios: (i) the default model, which has been fitted to the epidemiological data (community cases, hospital admissions, hospital occupancy and deaths, from the start of the pandemic until June 2022), replaying the recorded vaccination times (corresponding to 12-week and then 8-week intervals); (ii) a model in which the vaccination order is reversed, but the interval is fixed at 12-weeks, such that the youngest adults, most responsible for transmission, are vaccinated first; (iii) a model in which the order of receiving the first dose is preserved but second doses are given after a 3-week interval, with all efficacy parameters the same as with the 12-week interval; and finally (iv) a model with a 3-week dose interval and the same deployment as model (iii) that also captures the potential vaccine efficacy differences of a shorter dose interval. In all four scenarios, the number of doses administered each day remains as observed, as does the finally attained uptake within each age-group. We consider hospital admissions and deaths from December 2020 (when the vaccine campaign began) to September 2021 (when booster doses started), and assume that all other policy decisions (such as the steps taken as part of the Roadmap out of lockdown [16]) and population behaviour are unaffected by the scenario chosen.

To account for the vaccine efficacy differences arising from a shorter interval, we rely on the analyses of Khoury *et al*. [17], which links the level of neutralising antibodies to the degree of vaccine protection, and the experimental work of the Com-COV (Comparing COVID-19 Vaccine Schedule Combinations) group [18], which measured neutralising antibodies for 4 and 12 week dose intervals. Comparing a 4-week interval to the longer 12-week interval [18] reports a 2.35 (1.75-3.16) reduction in pseudotyped virus neutralised antibody level for two doses of the AstraZeneca ChAdOx1 vaccine, and a 1.44 (1.14-1.81) reduction for two doses of the Pfizer BNT162b2 vaccine. Using the relationship in [17], these lower neutralisation results for the shorter interval can be used to determine the vaccine efficacy against symptomatic infection following a second dose. Against the Alpha variant, the ChAdOx1 efficacy is reduced from 80% to 66% while the BNT162b2 efficacy is reduced from 88% to 82%; similar reductions from 70% to 49% and 90% to 85% are obtained against Delta variant. Given the lack of evidence, we assume that efficacy against severe disease (hospital admission or death) and the rate of waning protection is unaffected by the dose interval; as such, our results are possibly a best-case scenario.

Our results show that the policy of vaccinating the oldest (and most vulnerable) first (Fig. 1a,b black line) led to a faster decline in hospital admissions and deaths than targeting the younger age groups that are more responsible for transmission (Fig. 1a,b green line). However, a youngest first policy leads to lower hospital admissions (but not deaths) during the subsequent Delta wave (after July 2021) due to the later vaccination of older age groups together with greater population level immunity providing greater protection in the wider range of ages most likely to require hospital treatment. There are fewer early differences when keeping the prioritisation order but reducing the dose interval (Fig. 1 red and dashed pink lines), but there is still a cumulative net benefit in following the default scenario (Fig. 1c,d). In particular, by September 2021, a 3-week dose interval is predicted to lead to 38,400 (95% prediction intervals (PI): 31,900-46,600) more hospital admissions (Fig. 1c) and 4,600 (95% PI: 3,700-5,500) more deaths (Fig. 1d) than with the intervals deployed. These adverse effects increase to 56,600 (95% PI: 45,800-71,500) additional hospital admissions and 7,400 (95% PI: 5,600-9,400) deaths when assuming the lower efficacy against infection associated with a 3-week interval, and values would be even higher if we had assumed that the efficacy against severe disease was reduced by the shorter interval (Table 1). This echoes model-based findings that have been shown for other European countries [19, 20], although here we use a model that more robustly captures the temporal dynamics.

**Table 1:** Deaths and hospital admissions in England from 8th December 2020 to 1st September 2021, as observed (top row) and from four model scenarios as described in the main text - giving the mean and 95% prediction intervals. It should be stressed that all model results assume the same relaxation of control measures throughout 2021. All model results are from 400 samples of the posterior distribution.

|  | Deaths | Hospital Admissions |
| --- | --- | --- |
| Observed | 62163 | 243573 |
| (i) Default | 61,200 (59,900-62,400) | 245,800 (243,500-247,900) |
| (ii) Prioritise youngest | 84,500 (71,200-163,600) | 261,400 (223,600-487,100) |
| (iii) 3-week interval, default efficacy | 69,300 (67,000-71,100) | 314,100 (302,800-329,300) |
| (iv) 3-week interval, lower efficacy | 74,900 (70,700-79,900) | 341,400 (322,400-368,600) |

**Fig. 1:**
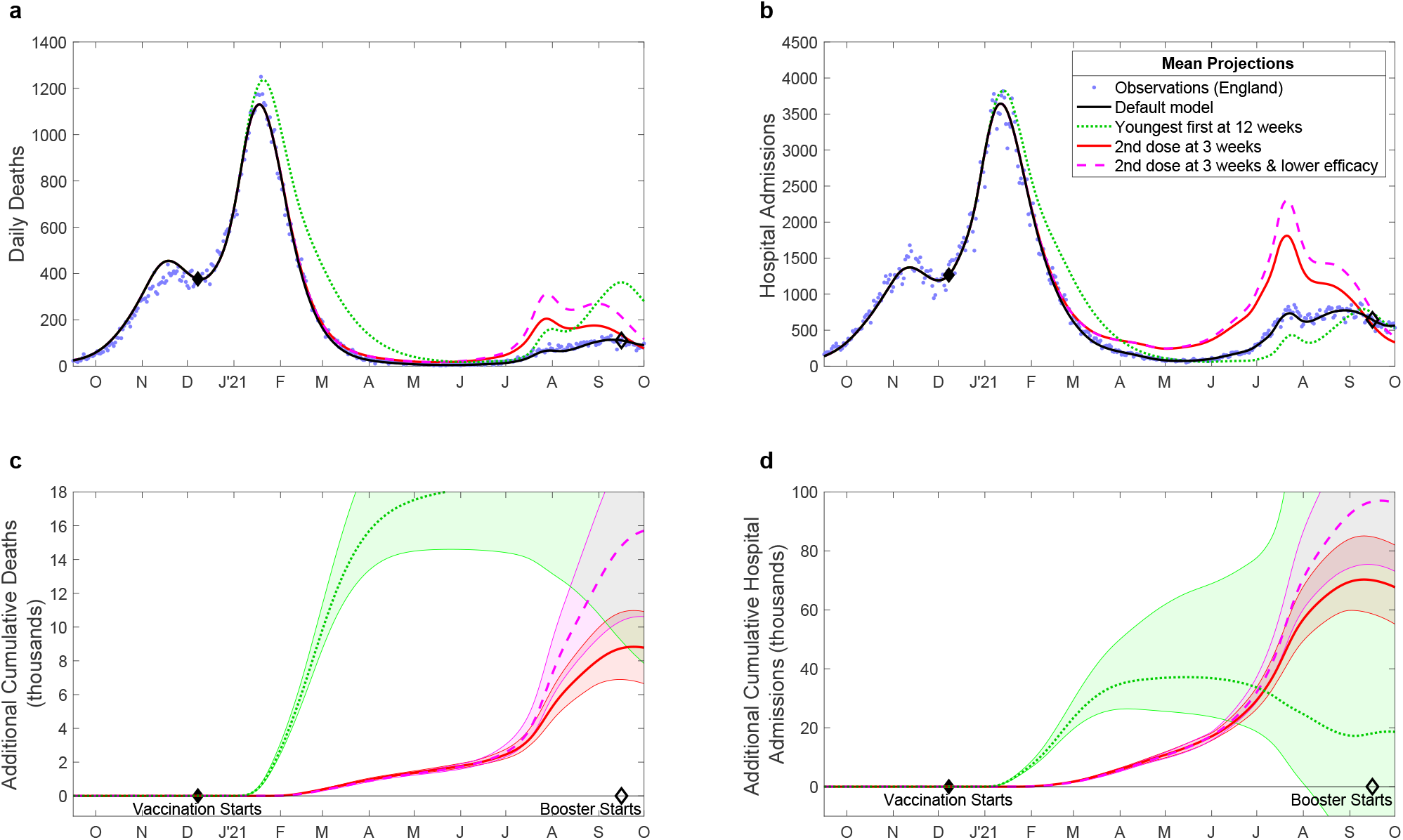
Projected changes in daily deaths (**a, c**) and hospital admissions (**b**,**d**) with differing vaccination patterns. (**a, b**) Compares the observed levels of severe outcomes (deaths or hospital admissions, blue dots) with (i) the default model projection using the recorded pattern of vaccination (black solid line) and alternative models where this pattern is perturbed: (ii) the ordering of vaccination is reversed so that younger adults are vaccinated first (green dotted line); (iii) vaccinations are given 3 weeks apart although the total number of daily doses administered is unaltered (red solid line); and (iv) the same 3-week strategy as the red line, but accounting for lower efficacy from the shorter dose interval (pink dashed line). (**c, d**) The cumulative additional deaths (**c**) and hospital admissions (**d**) projected from alternative assumptions, showing 95% prediction intervals (shaded regions) as well as mean values (lines). All results are from model simulations using 400 samples from the parameter posterior distributions.

As an extension to these results, we consider in more detail the impact that the age-dependent order of vaccination has on the projected number of deaths and hospital admissions due to COVID-19. We chose a random sample of 10,000 different age-orders, while maintaining the number of doses delivered per day and the recorded uptake in each age-group. We note that this is an idealised scenario in which vaccination within each age-group is completed (based on their recorded uptake) before the next age-group is vaccinated, and therefore cannot be closely compared to the realised order. We report the number of hospital admissions and deaths during two periods: the Alpha variant wave after the start of the vaccination campaign (from 8th December 2020 to 15th May 2021) (Fig. 2 x-axis) and for the entire period from the start of vaccination to boosters (8th December 2020 to 1st September 2021) (Fig. 2 y-axis). For deaths across both time periods, and for hospital admissions in the Alpha wave, the strategy of vaccinating in age-order from oldest to youngest appears close to optimal, re-enforcing the conclusions from earlier work on vaccine prioritisation [4, 21]. When considering total hospital admissions over the longer time scales however, a simple policy of oldest first is sub-optimal - strategies that initially target those over 80 then try to generate broad levels of immunity in the community perform better.

**Fig. 2:**
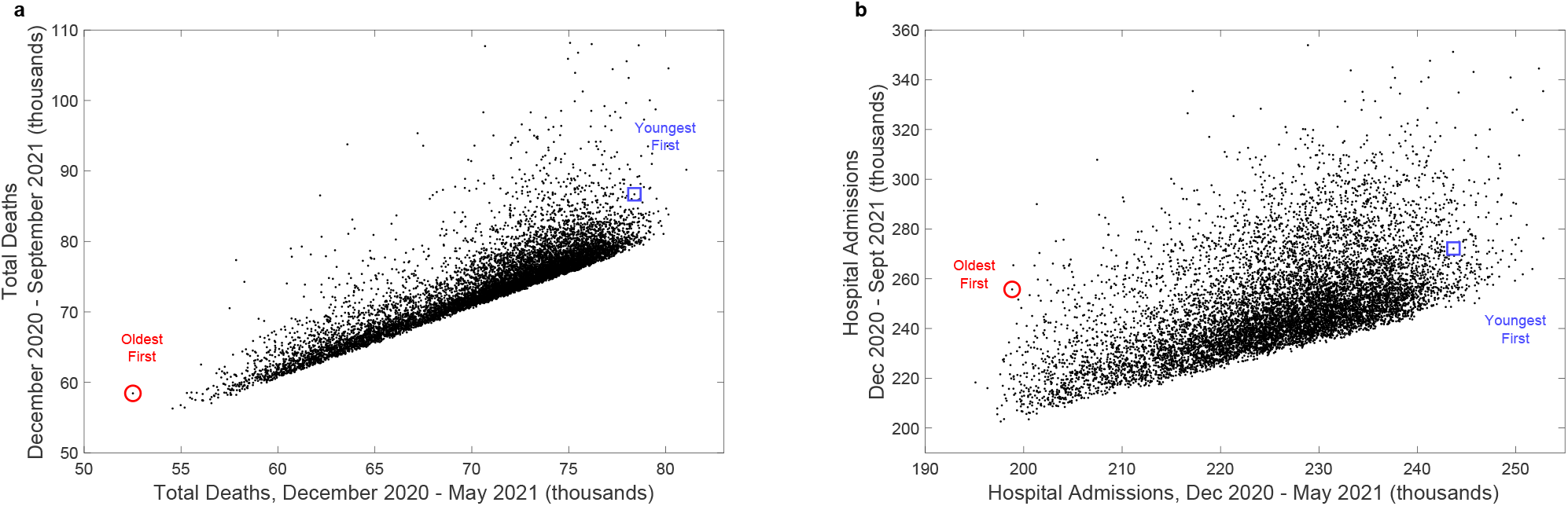
Prediction of deaths and hospital admissions for a range of age-dependent vaccination priority orders. For 10,000 randomly chosen age-based priority orders the black dots show the total expected number of deaths (**a**) and hospital admissions (**b**) over the remainder of the Alpha wave (8th December 2020 to 15th May 2021, x-axis) and over the period until boosters (8^th^ December 2020 to 1st September 2021, y-axis). We mark the results associated with vaccinating in chronological order oldest first (red circle) and youngest first (blue square).

## Discussion

A key question when planning an emergency vaccination programme against an epidemic/pandemic is how to optimally target the vaccine. One argument is that those most vulnerable to severe disease outcomes should be prioritised in order to reduce hospital admissions and deaths [4, 22], even if this is sub-optimal for reducing the levels of infection. An alternative argument is that the vaccine should be targeted at those most responsible for driving transmission [3], thereby limiting the level of infection and as a consequence reducing severe disease. We note that the first strategy only depends on a high vaccine efficacy against severe disease, while the second relies on a high efficacy against infection and onward transmission.

Our model-based approach has shown that the adopted strategy of targeting the elderly and vulnerable is close to the optimal, especially when considering COVID-19 mortality. We attribute this to the high level of infection when the vaccination programme began, such that limiting severe disease by targeting younger high-transmitters takes too long to become an effective strategy. Targeting large numbers of high transmitters could be effective when case numbers are low, suggesting a complex interdependence between robust long-term projections and determining the optimal prioritisation strategy. The adopted strategy in the UK had the additional advantage that it was simple to explain and implement. When considering hospital admissions over both the Alpha and Delta waves, other strategies become optimal - in general these target early doses at the over 80s (generating protection in this most vulnerable age-group) before switching to younger age-groups in a bid to minimise the reproductive ratio of the invading Delta variant – echoing the idea that it may be better to target high-transmitters during periods of low infection. While such optimal strategies can also perform well in terms of reducing mortality, they could not have been determined without knowledge of the entire epidemic trajectory and lack the operational simplicity of the adopted strategy.

A similar question arises when considering how to prioritise the two doses of vaccine: is it better to generate high immunity in a few individuals by giving them two doses with a short separation interval, or to generate weaker immunity in twice as many by increasing the dose interval. The optimal choice depends on the heterogeneity in risk [9], the time-varying force of infection and the immunological responses to first and second dose at varying intervals. Here we have shown that a longer interval, providing more first doses as early as possible, is optimal; and the benefits are even greater when the higher levels of neutralising antibodies from 12-week interval [18] is taken into account.

While these results are compelling, there are some limitations to the method. Conceptually, we have assumed that the same behavioural changes (including population precautionary behaviour and age-dependent vaccine uptake) and legal changes (such as the relaxation steps out of lockdown) would occur irrespective of the vaccination scenario; this is unlikely to be true as the higher number of hospital admissions and deaths observed in some scenarios (especially if vaccinating younger individuals first) would likely shift the relaxation of restrictions and illicit a greater precautionary reaction from the general public [16]. Our model is age-structured and parameterised to fit the observed epidemic, but does not partition the population by other risk factors (primarily because most of the epidemiological data is not partitioned by additional factors such as comorbidities). Therefore, the model cannot fully capture the prioritisation of vulnerable risk groups, especially when these are in younger ages. Longer-term dynamics, beyond September 2021 and the start of the booster programme, become more difficult to predict and would be contingent on assumptions about how the boosters were deployed and the immune landscape, which is more likely driven by infection rather than vaccination in the non-default scenarios.

With the advantage of hindsight, the JCVI’s advice to prioritise first doses of SARS-CoV-2 vaccine to as many elderly or vulnerable individuals as possible early in 2021 appears to have been extremely well judged and has substantially reduced the public health burden of severe COVID-19 in England. In general, we would also expect such results to hold for future pandemics in other geographical or demographic settings whenever first doses generate substantial protection against severe disease in the most vulnerable [9]. However, bespoke models matched to available national data are ideally needed to assess the benefits on a case-by-case basis [20, 22, 23].

Public health decisions often have to be made on the basis of limited data and therefore need to be as robust as possible to potential changes in scientific understanding. The advice given by JCVI in late 2020 and early 2021, was based on very limited data about vaccine efficacy against the newly emerged Alpha variant, and with no knowledge of the forthcoming waves of infection or the waning of protection. Under such conditions a simple yet precautionary strategy is optimal, rapidly providing broad protection to those individuals that need it the most.

## Methods

Here we detail the underlying mathematical framework that defines the transmission model. We break the model description into multiple sections that combine to generate a picture of SARS-CoV-2 transmission and COVID-19 disease burden in England. The model structure has also been detailed in previous publications [4, 16, 21, 24, 25].

## Model overview

The model is built around the traditional deterministic SEIR (Susceptible, Exposed, Infectious, Recovered) framework [26], with three exposed classes to capture the distribution of times from infection to becoming infectious [27], and splitting the infectious group into symptomatic and asymptomatic infection [24]. To this simple model we add additional structure to capture the effects of restricted social interaction whilst maintaining household transmission [24]. We then ‘replicated’ this fundamental model twenty-one times to mimic five-year age-groups (0 − 4, 5 − 9, …, 100+), and a further seven times to capture the different dynamics in the seven National Health Service (NHS) regions in England. Collectively, the model is written as a large system of ODEs (ordinary differential equations, as detailed in the Supplementary Information).

This basic model was sufficient for the early waves of infection (from January to November 2020), comprising a single variant without vaccination and without large amounts of reinfection. During this early phase of the pandemic, the main driving parameter was the level of precautionary behaviour in the population, which determined the level of social-mixing and therefore the scale of transmission outside the household [16, 24], although we also fitted a number of other parameters (including case:hospitalisation and case:mortality ratios, age-dependent effects and the relative strength of asymptomatic compared to symptomatic transmission) [28]. From the age-structured symptomatic class, we calculate the number of severe health episode outcomes (hospital admissions, intensive care unit admissions and deaths), these being key public health observables and measures of concern for this pandemic, although these quantities do not impact the transmission dynamics (Fig. 3). We performed the fitting in a Bayesian framework, matching the data on the daily hospital admissions, hospital occupancy, ICU occupancy, deaths and proportion of community (Pillar 2) tests that are positive in each of the seven NHS regions of England to a Poisson distribution with a mean given by the ODE model [28].

**Fig. 3:**
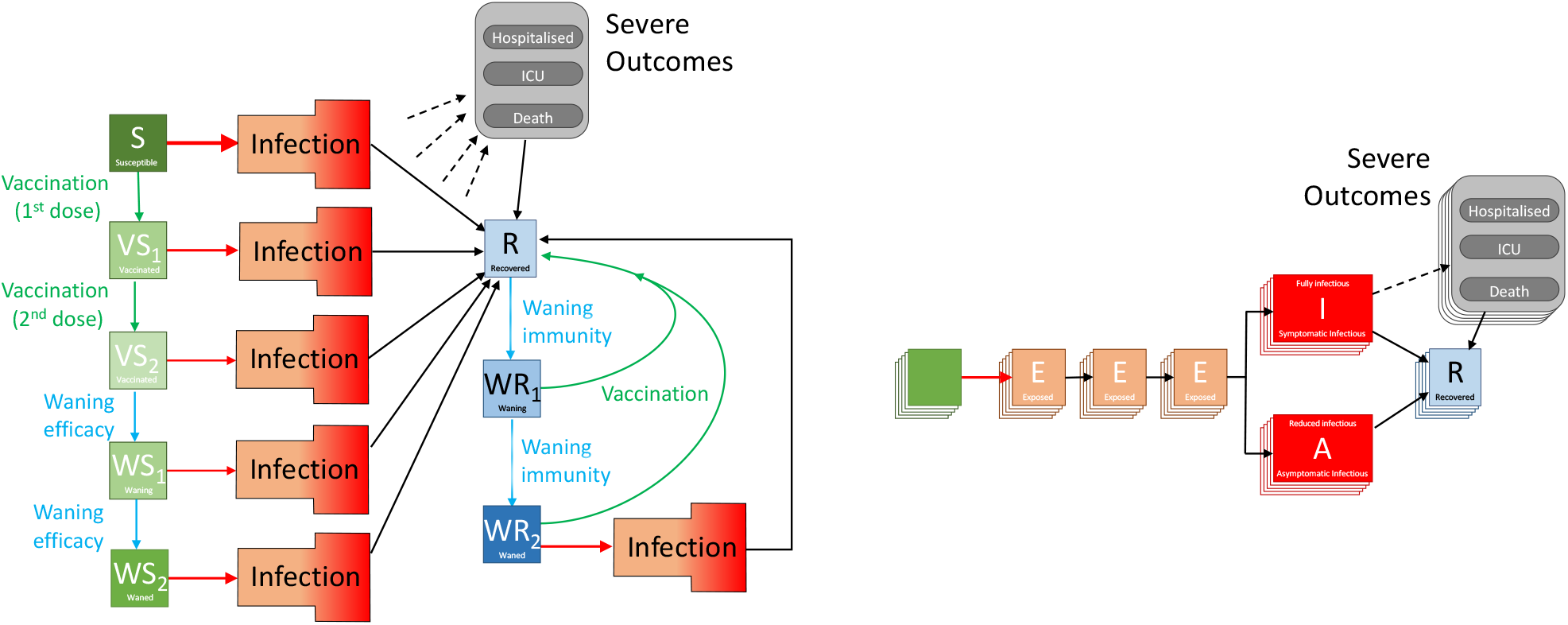
Graphical schematic of the models used in the paper. The left-hand schematic shows the progress between different compartments. Infection is shown as a red arrow (with the size providing some indication of the risk), recovering is shown by black arrows, the two-dose vaccination is shown in green, while waning protection is in light blue. The infection compartment is multi-dimensional and shown in the right-hand schematic. It captures infection, the multiple latent classes and whether the infectious class is associated with symptomatic or asymptomatic infection. Each compartment is shown as multi-layered to indicate additional age-, regional- and variant-specific heterogeneity. Severe outcomes of hospital or ICU admission and death (shown with dashed arrows) are related to the level of infection in each age-group and their degree of vaccine (or infection-induced) protection.

From late 2020, variants increased the dimensionality of this model. Each new variant required a duplicate of all the infected model classes to capture differences in transmission and risks of severe outcomes; the rise of each variant was captured by additionally fitting to the proportion of S-gene target failures (a measure of variant-type) from TaqPath PCR testing [29, 30]. Later iterations of the models incorporated the multiple Omicron variants that have caused major waves in the UK, and included reinfection - assuming that infection with a previous variant only induced partial cross-immunity. For the Alpha and Delta variant model used in this work, we assumed full cross-immunity, which is in close agreement with the observation during this time period that only 1% of reported cases had previously reported infection [31].

The start of the vaccination campaign in December 2020 necessitated a further partitioning of the population by vaccination status, allowing us to capture both the reduced risk of infection and the reduced risk of severe outcomes. Early models used in the initial assessment of vaccine dose intervals only considered wildtype and Alpha variants [9], although the work presented here also includes the Delta variant, with different levels of vaccine protection for each variant. It became clear during 2021 that the vaccines (and to a less extent infection) did not confer long-lasting immunity [32] and therefore waning levels of protection both in terms of vaccine-induced and infection-induced immunity were added (generating additional elements within the model). This is shown schematically in Fig. 3, focusing on vaccination and waning immunity. For simplicity the structure within the infection process is not fully represented (which includes exposed and infectious classes as well as variant structure); the full model is also partitioned into five year age groups and seven different spatial regions, with parameters fitted to the dynamics in each of these regions (although not to age-structured data). The most recent versions of the model additionally include the action of booster vaccination, which resets protection but is itself subject to waning, although this is excluded in this description framework which only considers the period before Autumn 2021 boosters were available.

More information of the individual components that combine to generate the full model are given below, with the associated equations and mathematical details provided in the Supplementary Information. First we consider the two aspects that are key to this work: vaccination modelling and the impact of the dose-interval on protection. Subsequent subsections offer expanded details on our methodological approach and assumptions for the infection model, age and transmission structure, quarantining and isolation, regional modelling, variant modelling and, lastly, parameter inference.

### Vaccination Modelling

We capture vaccination using a leaky approach, such that all individuals that have been vaccinated have a reduced susceptibility to infection and a subsequently lower risk of symptoms or severe disease. However, earlier versions of our model that made the alternative non-leaky (all-or-nothing) assumption [4, 21] produce similar results, echoing the insensitivity found by other researchers [22].

The model captures three important aspects of vaccination and protection:

Firstly, individuals are given two doses of vaccine with the first conferring only partial immunity (see Table 2). Vaccination is age-dependent but is independent of infectious state such that both susceptible and recovered individuals are vaccinated, although each person can only receive one course of vaccine. In this default model we replay the recorded pattern of vaccination, giving recorded first doses to individuals that have not yet been vaccinated and giving recorded second doses to those that have only received their first dose.

**Table 2:**
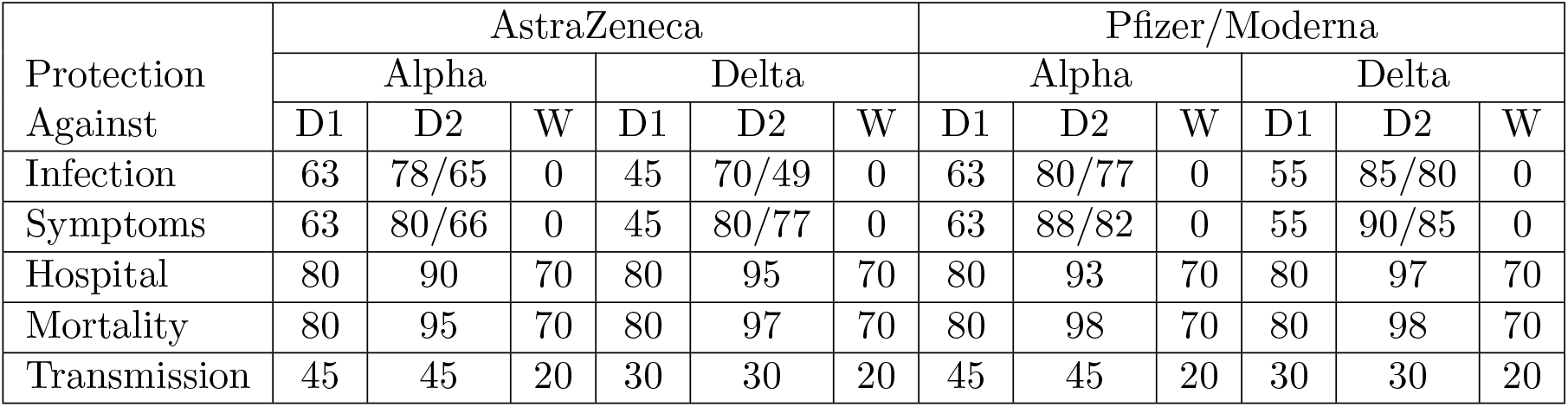
Assumed percentage protection from the two vaccine types against the Alpha and Delta variants after one dose (D1), after two doses (D2, shown for the dose interval gaps of 12-weeks/3-weeks), and after waning (W). These values correspond to relatively slow waning of vaccine protection (*ω*_1_ = 100^*−*1^ per day, *ω*_2_ = 320^*−*1^ per day, 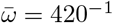 per day) [33–35].

| Protection<br>Against | AstraZeneca |  |  |  |  |  | Pfizer/Moderna |  |  |  |  |  |
| --- | --- | --- | --- | --- | --- | --- | --- | --- | --- | --- | --- | --- |
|  | Alpha |  |  | Delta |  |  | Alpha |  |  | Delta |  |  |
|  | D1 | D2 | W | D1 | D2 | W | D1 | D2 | W | D1 | D2 | W |
| Infection | 63 | 78/65 | 0 | 45 | 70/49 | 0 | 63 | 80/77 | 0 | 55 | 85/80 | 0 |
| Symptoms | 63 | 80/66 | 0 | 45 | 80/77 | 0 | 63 | 88/82 | 0 | 55 | 90/85 | 0 |
| Hospital | 80 | 90 | 70 | 80 | 95 | 70 | 80 | 93 | 70 | 80 | 97 | 70 |
| Mortality | 80 | 95 | 70 | 80 | 97 | 70 | 80 | 98 | 70 | 80 | 98 | 70 |
| Transmission | 45 | 45 | 20 | 30 | 30 | 20 | 45 | 45 | 20 | 30 | 30 | 20 |

Secondly, we assume that immunity from either vaccination or infection wanes over time. This is captured in a two-step (two-compartment) process, first moving individuals into an initial compartment where protection is maintained before subsequently moving to the final waned compartment with lower (but non-zero) levels of protection. This two-step process enables us to better capture the observed time-scales for loss in protection (Fig. 4). Waning from the recovered state after infection is assumed to progress more slowly than following vaccination.

**Fig. 4:**
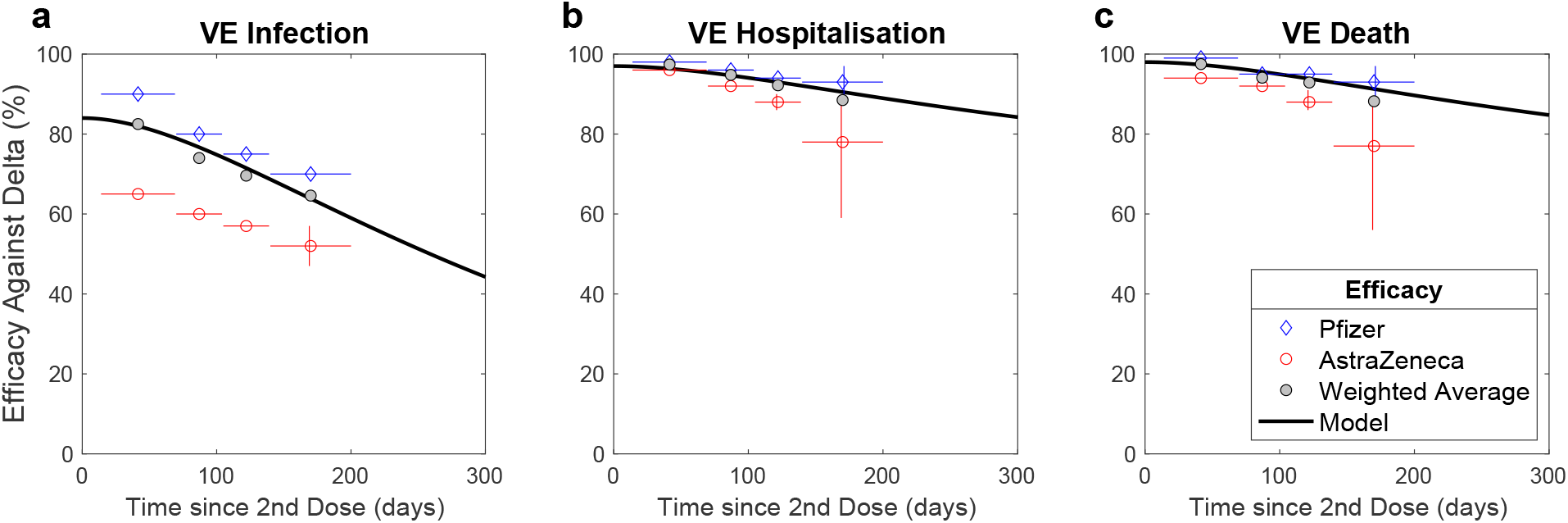
Comparison between model assumptions and estimates of vaccine efficacy against the Delta variant for infection (a), hospitalisation (b) and death (c). Data are taken from UKHSA estimates in December 2021 [33]; data is shown for the Pfizer vaccine (blue diamonds), AstraZeneca vaccine (red circles) and the weighted average (grey), the model obeys the simple compartmental rules described above. The vertical lines represent the 95% confidence intervals, capturing uncertainty in the data; while the horizontal lines correspond to the period over which the data is amalgamated.

Finally, the degree of protection (vaccine efficacy) is dependent on the number of doses received, whether or not protection has waned, which vaccine was used (AstraZeneca or Pfizer/Moderna) and the variant involved (Table 2). Additionally, vaccine-derived protection differs, with protection against infection being the weakest and protection against mortality being the strongest. As such an individual vaccinated with two doses of Pfizer, with a dose interval of 12 weeks, may have 85% protection against infection (that is their risk of infection is reduced to 15% compared to an unvaccinated person of the same age) while protection against mortality is 98% (their risk is reduced to just 2% compared to an unvaccinated person, although this includes the reduced risk of being infected). Protection in the waned state remains relatively high against severe disease but protection against infection is assumed to drop to zero; this leads to a rapid decline in vaccine efficacy against infection but slower declines against hospitalisation or death in agreement with available data (Fig. 4). All these values are taken from a range of studies by PHE or UKHSA [33], that were conducted throughout the pandemic, with reports published since May 2021; the values are in close agreement with aggregate estimates agreed by the UK SAGE subcommittee [34].

While for the default model we can simply replay the number of first and second doses of vaccine given to each age-group, for the other counterfactual scenarios we need to generate these synthetically.

For model (ii) where we prioritise the youngest first, rather than the elderly first, we initially generate a list of first doses by age group which is reversed to give the new order in which first doses are delivered. Each day, we use the total number of observed doses (both first and second) to define the regional capacity. From this capacity, second doses are prioritised such that individuals get their second dose at approximately 12-weeks after receiving their first dose; any spare capacity is used to give first doses working through the ordered list.

For models (iii) and (iv) we maintain the recorded order of the first-doses. As with model (ii) the regional capacity is determined by the historic pattern of vaccination. The capacity is first used to give second doses at 3-weeks after the first dose, with spare capacity used to work through the ordered list of those wanting a first doses. The only difference between models (iii) and (iv) are the assumptions about different levels of protection against infection and symptoms.

By following this procedure, we preserve both the daily deployment capacity and the uptake within each age-group, which are the same for all four models.

### Impact of Shorter Dose-Interval on Vaccine Efficacy

When focusing on the impact of a shorter dose-interval, we considered a scenario where vaccine efficacy remained unaffected and a scenario in which efficacy was reduced. For the scenario where a shorter dose-interval resulted in a reduced efficacy, the reduction in efficacy was guided by two studies: (i) the analyses of Khoury *et al*. [17] (Fig. 5 left-hand panel), which linked the level of neutralising antibodies to the degree of vaccine protection against symptomatic disease; (ii) the experimental work of the Com-COV group [18], which highlighted a shorter interval between vaccine doses being associated with a lower level of neutralising antibodies. We took efficacy estimates after the second dose using 12-week intervals from UKHSA studies [33] and translated via a three-step process into the reduced efficacy at 3 or 4 weeks (details in Fig. 5 central panel). The resultant values are shown in the right-hand panel of Fig. 5 and are given in Table 2.

**Fig. 5:**
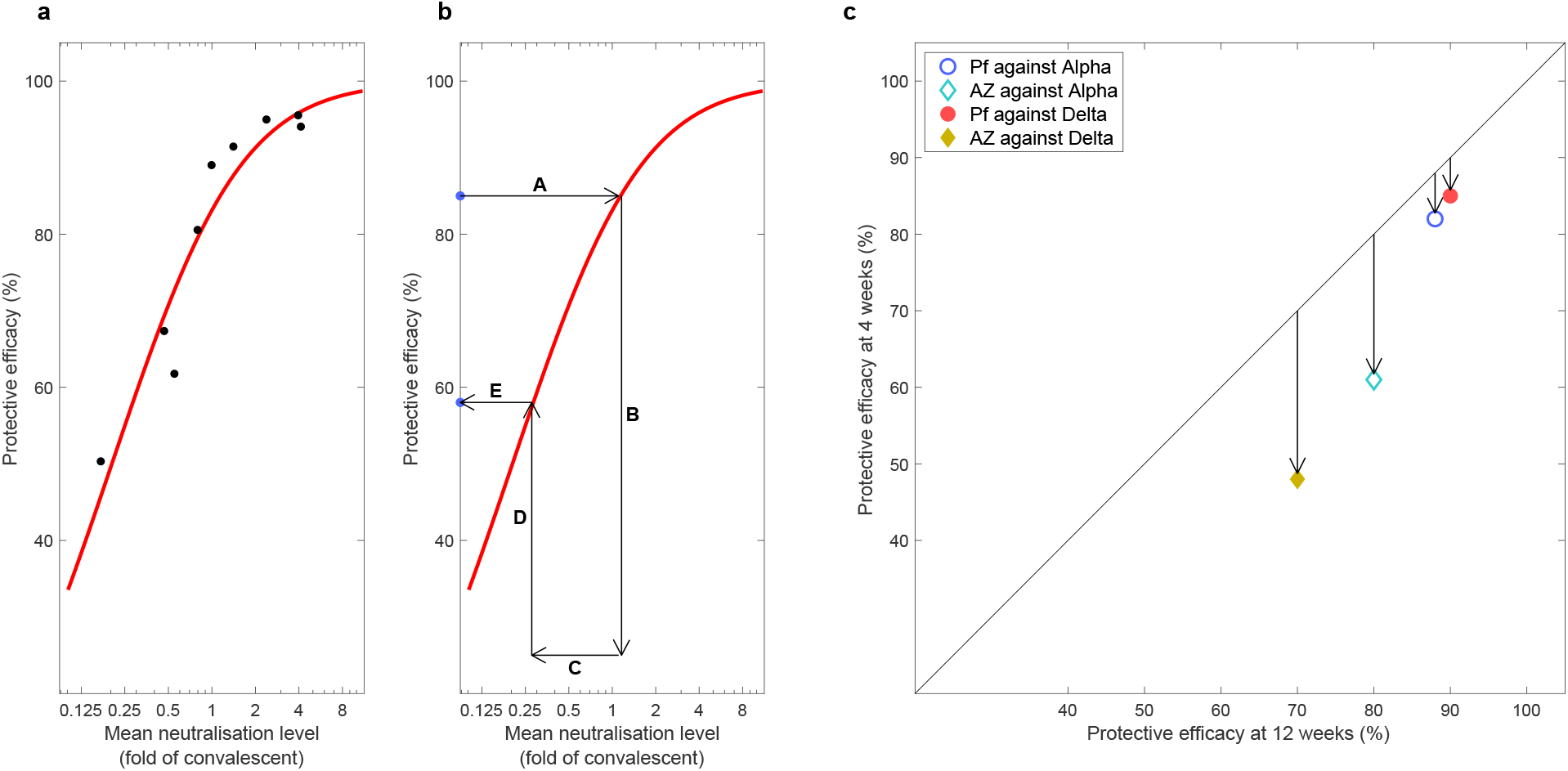
Calculation of the impact of shorter dose-interval on vaccine efficacy. (**a**) The data (dots) and fitted curve (red line) from [17] that translates neutralisation level (relative to that for convalescent) to the degree of protection against symptomatic disease. (**b**) The steps needed to scale efficacy for a 12-week dose interval to that for a short 3- or 4-week dose interval: (steps A-B) 12-week efficacy is translated to a relative neutralisation level via the red curve [17]; (step C) neutralisation level is reduced according to the values obtained by the Com-COV study [18]; (steps D-E) the reduced neutralisation level is translated to a reduced efficacy for the shorter dose interval. (**c**) The consequences of this process for the AstraZeneca (AZ) and Pfizer (PF) vaccines against the Alpha and Delta variants, and hence the modified vaccine efficacy estimates for model (iv).

### Infection modelling

As is common to most epidemiological modelling we stratify the population into multiple disjoint compartments and capture the flow of the population between compartments in terms of ordinary differential equations. At the heart of the model is a modified SEIR equation, where individuals may be susceptible (*S*), exposed (*E*), infectious with symptoms (*I*), infectious and either asymptomatic or with very mild symptoms (*A*) or recovered (*R*). Both symptomatic and asymptomatic individuals are able to transmit infection, but asymptomatic infections do so at a reduced rate given by *τ*. Hence, the force of infection is proportional to *I* + *τ A*. The separation into symptomatic (*I*) and asymptomatic (*A*) states within the model is somewhat artificial as there are a wide spectrum of symptom severities that can be experienced [36], with the classification of symptoms changing over time. Our classification reflects early case detection, when only relatively severe symptoms were recognised.

To obtain a better match to the time from infection to becoming infectious, we model the exposed class as a three-stage process, such that in a stochastic formulation the distribution of the latent period would be an Erlang distribution.

### Age Structure and Transmission Structure

We expanded the simple SEIR-type model structure to divide each compartment into twenty-one five-year age-groups (0-4, 5-9, …., 95-99, 100+). Based on observations of the epidemic, age has three major impacts on the epidemiological dynamics, with each element parameterised from the available data. Firstly, and most notably, older individuals who become symptomatically infected have a higher risk of more severe consequences of infection, including hospital admission and death [9]. Secondly, older individuals have a higher susceptibility to SARS-CoV-2 infection. Finally, older individuals have a higher risk of developing symptoms, and therefore have a greater rate of transmission per contact.

One of the key components of age-structured models is the contact and hence risk of transmission between age-groups. Contacts are captured by age-structured matrices, with transmission then modified by age-dependent susceptibility. We consider contacts in four settings: household (***β***^*H*^), school (***β***^*S*^), workplace (***β***^*W*^) and other settings (***β***^*O*^); these matrices are combined with the age-dependent susceptibility and transmissibility vectors to generate a force of infection. We took these matrices from Prem et al. [37] to allow easy translation to other geographic settings, although other sources such as the POLYMOD matrices [38] or estimates produced during the pandemic, such as from the CoMiX study [39], could be used.

One of the main modifiers of mixing and therefore transmission is the level of precautionary behaviour, *ϕ* [16]. This time-varying scaling parameter is used to modify the contact matrices, such that when *ϕ* = 1 mixing in workplaces and other settings take their lowest value, whereas when *ϕ* = 0 the mixing returns to pre-pandemic levels [24]. A high *ϕ* value also corresponds to an increase in household mixing on the assumption that more time will be spent within the household environment. Mixing within the school setting followed the prescribed opening and closing of schools, capturing both the normal pattern of school holidays as well as the closing of schools at times of peak infection (March-July 2020, January-March 2021). The weekly values of *ϕ* are inferred by fitting the model to epidemic data, producing trends that are in close agreement with Google mobility [40] and CoMix [39] data sets that track the changes in movement and mixing over the pandemic (see Supplementary Information). It should be noted however, that *ϕ* provides a more nuanced estimate than either of these primary sources, as it also seeks to capture the behaviour of individuals when they mix - including the propensity of infected individuals to test and isolate.

To ensure that we can replicate the long-term dynamics of infection we allow the population to age. The ageing process occurs annually (corresponding to the new school year in September) in which approximately one fifth of each age-group moves to the next oldest age cohort — small changes to the proportion moving between age-groups are made to keep the total population size within each age-group constant.

### Quarantining and Isolation

One of the key characteristics of the COVID-19 pandemic in the UK has been the use of self-isolation and household quarantining to reduce transmission. We approximate this process by distinguishing between first infections (caused by infection related to any non-household mixing) and subsequent household infections (caused by infection due to household mixing). The first symptomatic case within a household (which might not be the first infection) has a probability (*H*) of leading to household quarantining; this curtails the non-household mixing of the individual and all subsequent infections generated by this individual. This formation has been shown to be able to reduce the reproductive ratio, *R*, below one even when there is strong within household transmission (see Supplementary Information), as infection from quarantined individuals cannot escape the household [24].

### Regional Modelling

The model operates at the scale of seven NHS regions within England (East of England, London, Midlands, North East, North West, South East and South West). For simplicity and speed of simulation we assume that each of these regions acts independently and in isolation - we do not model the movement of people or infection across borders. In addition, the majority of parameters are regionally specific, reflecting different demographics, deprivation and social structures within each region. However, we include a hyper-prior on the shared parameters such that the behaviour of each region helps inform the value in others.

### Variant Modelling

The model also captures the main SARS-CoV-2 variants that have been responsible for most infections in England up to September 2021: the wildtype virus (encapsulating all pre-Alpha variants), the Alpha variant and the Delta variant. Each of these requires a replication of the infectious states for each variant type modelled. We assume during this period that infection with each variant confers immunity to all other variants, such that there is indirect competition for susceptible individuals. This assumption is relaxed in later iterations of the model which include the Omicron variant, but for the wildtype, Alpha and Delta waves, only 1% of reported cases were reinfections [31] suggesting this is a justifiable simplification.

The competition and eventual dominance of each variant is driven by the transmission advantage, estimated by matching to the proportion of positive community PCR tests (Pillar 2 test) that are positive for the S-gene. The TaqPath system that is used for the majority of PCR tests in England is unable to detect the S-gene in Alpha variants (due to mutations in the S-gene). The switch from S-gene positive to S-gene negative and back to S-gene positive corresponds with the dominance of wildtype, Alpha and Delta variants, respectively. We infer the transmissibility of Alpha to be 52% (CI 35-71%) greater than wildtype, and the transmissibility of Delta to be 71% (CI 52-107%) greater than Alpha.

### Parameter Inference

Key to the accuracy of any model are the parameters that underpin the dynamics. With a model of this complexity, a large number of parameters are required. Some, such as vaccine efficacy, are assumed values based on the current literature; while others are inferred from the epidemic dynamics.

Of the inferred parameters there are three basic classes; those, such as scalings of the case-hospitalisation ratios, that are different between regions and variants; others such as age-dependent susceptiblity that are universal (the same for all regions and variants); and finally the level of precautionary behaviour over time which changes on a weekly time-scale. Bayesian inference, using an MCMC process, is applied to each of the seven NHS regions in England to determine posterior distributions for each of the regional parameters (further details are given in [28] and the Supplementary Information). The distribution of parameters leads to uncertainty in model projections, which is represented by the 95% prediction interval in all graphs (this is the interval that contains 95% of all predictions). We note that when we compare two scenarios (for example vaccination with a 3-week interval, with vaccination using a 12-week interval) we compare simulations with the same parameters chosen from the posterior distributions – and then calculate means and 95% prediction intervals based on these results.

As the epidemic has progressed, new posterior distributions based on the latest data are initialised from previous MCMC chains – ensuring a rapid fit to historical data. In general this refitting process has been performed weekly (or twice weekly) throughout the pandemic. For the time period of relevance in this paper (December 2020 - September 2021), we matched to six observations: hospital admissions, hospital occupancy, ICU occupancy, deaths, proportion of pillar 2 (community) test that are positive, and the proportion of pillar 2 tests that are S-gene positive (as a signal of the ratio of wild-type to Alpha variant, then a signal of the ratio of Delta variant to Alpha variant). We note that in [28], which was written in the early stages of the pandemic, we did not fit to S-gene data as we had been dealing with a single variant. Although not part of the underlying transmission dynamics, these six quantities for each spatial region can be generated from the number, age and type of infection within the model. We compared observations and model generated results by considering the likelihood of generating the observations when assuming them to be Poisson distributed (for numbers) or binomially distributed (for proportions), with the means of these distributions given by the results of the deterministic model.

## Data Availability

Data on cases were obtained from the COVID-19 Hospitalisation in England Surveillance System (CHESS) data set that collects detailed data on patients infected with COVID-19. Data on COVID-19 deaths were obtained from Public Health England. These raw data contain confidential information, with public data deposition non-permissible for privacy reasons. The CHESS data resides with the National Health Service (www.nhs.gov.uk) whilst the death data are available from Public Health England (www.phe.gov.uk). Again these raw data contain confidential information, with public data deposition non-permissible for privacy reasons. The ethics of the use of these data for these purposes was agreed by Public Health England with the Governments SPI-M-O / SAGE committees. Processed data (which is more aggregated) is freely available from the UK Coronavirus dashboard: https://coronavirus.data.gov.uk/

## Code Availability

Specific code relating to this paper will be available shortly. Generic code used to model COVID-19 in England is available at: https://github.com/MattKeeling/Dose_Interval.git

## Ethical Considerations

Data from the CHESS and SARI databases were supplied after anonymisation under strict data protection protocols agreed between the University of Warwick and Public Health England. The ethics of the use of these data for these purposes was agreed by Public Health England with the Government’s SPI-M-O / SAGE committees.

## Acknowledgements

MJK was supported through the JUNIPER modelling consortium [grant number MR/V038613/1]; MJK and SM were supported by the National Institute for Health Research (NIHR) [Policy Research Programme, Mathematical and Economic Modelling for Vaccination and Immunisation Evaluation, and Emergency Response; NIHR200411]; MJK and EMH were supported by the Medical Research Council through the COVID-19 Rapid Response Rolling Call [grant number MR/V009761/1] and the Biotechnology and Biological Sciences Research Council [grant number BB/S01750X/1]. MJK is affiliated to the National Institute for Health Research Health Protection Research Unit (NIHR HPRU) in Gastrointestinal Infections at University of Liverpool in partnership with UK Health Security Agency (UKHSA), in collaboration with University of Warwick. MJK is also affiliated to the National Institute for Health Research Health Protection Research Unit (NIHR HPRU) in Genomics and Enabling Data at University of Warwick in partnership with UK Health Security Agency (UKHSA). The views expressed are those of the author(s) and not necessarily those of the NHS, the NIHR, the Department of Health and Social Care or UK Health Security Agency.

## Author Contributions

**Matt J. Keeling**: Conceptualisation, Data curation, Formal analysis, Funding acquisition, Methodology, Software, Validation, Visualisation, Writing—Original draft, Writing—Review and Editing. **Samuel Moore**: Software, Validation, Visualisation, Writing—Review and Editing. **Bridget Penman**: Methodology, Writing—Review and Editing. **Edward M. Hill**: Software, Validation, Visualisation, Writing—Review and Editing.

