## Supplementary Information for "The Impact of SARS-CoV-2 Vaccine Dose Separation and Dose Targeting on Hospital Admissions and Deaths from COVID-19 in England"

Matt J. Keeling, Samuel Moore, Bridget Penman, Edward M. Hill

In this supplement we detail the underlying mathematical framework that defines the model dynamics for vaccination, waning, infection and severe disease. Although much of this is described in the Methods of the main paper, here we provide the differential equations that underpin the model and describe in more detail the fitting of the model to the available epidemiological data.

#### 1 Vaccination and Waning

In the absence of infection, there is still a complex pattern of vaccination and waning which is described here with the process of infection described below. The model replicates the action of:

- first and second doses of vaccine, at time-varying rates $v_1$ and $v_2$ respectively, that move susceptible individuals through to vaccinated states ($VS_1$ and $VS_2$) but have no impact on infected or recovered individuals - for model simplicity $v_1$ and $v_2$ correspond to the rate at which the impact of the vaccine dose takes effect, which is around 10 days post vaccination;
- waning vaccine efficacy at rates $\omega_1$ and $\omega_2$, giving a two-step process from fully vaccinated to waned efficacy - we also allow waning from state $VS_1$ at rate $\bar{\omega}$ (where $\bar{\omega}^{-1} = \omega_1^{-1} + \omega_2^{-1}$), although for those that receive two doses within 3 and 12 weeks this is uncommon;
- waning immunity from past infection at rates $\Omega_1$ and $\Omega_2$ which are assumed to be slower than the waning of vaccine efficacy.

The model also needs to capture the total number of individuals who have been given a first or second dose of vaccine ($V_1$ or $V_2$ out of a total population size of $N$) to ensure that only individuals that have not been vaccinated are offered a first dose, and only individuals that have been vaccinated once are offered a second dose.

Adding subscripts that signify age ($a$) and region ($r$), when concentrating on vaccination and waning immunity (i.e. ignoring infection and variants) the underlying equations become:

$$
\begin{aligned}
\frac{dS_{a,r}}{dt} &= -v_{1,a,r}\frac{S_{a,r}}{N_{a,r}-V_{1,a,r}} \\
\frac{dVS_{1,a,r}}{dt} &= v_{1,a,r}\frac{S_{a,r}}{N_{a,r}-V_{1,a,r}} - v_{2,a,r}\frac{VS_{1,a,r}}{V_{1,a,r}-V_{2,a,r}} - \bar{\omega}VS_{1,a,r} \\
\frac{dVS_{2,a,r}}{dt} &= v_{2,a,r}\frac{VS_{1,a,r}}{V_{1,a,r}-V_{2,a,r}} - \omega_1 VS_{2,a,r} \\
\frac{dWS_{1,a,r}}{dt} &= \omega_1 VS_{2,a,r} - \omega_2 WS_{1,a,r} \\
\frac{dWS_{2,a,r}}{dt} &= \omega_2 WS_{1,a,r} + \bar{\omega}VS_{1,a,r} \\
\frac{dR_{a,r}}{dt} &= -\Omega_1 R_{a,r} + v_{1,a,r}\frac{WR_{1,a,r}+WR_{2,a,r}}{N_{a,r}-V_{1,a,r}} \\
\frac{dWR_{1,a,r}}{dt} &= \Omega_1 R_{a,r} - \Omega_2 WR_{1,a,r} - v_{1,a,r}\frac{WR_{1,a,r}}{N_{a,r}-V_{1,a,r}} \\
\frac{dWR_{2,a,r}}{dt} &= \Omega_2 WR_{1,a,r} - v_{1,a,r}\frac{WR_{2,a,r}}{N_{a,r}-V_{1,a,r}} \\
V_{1,a,r}(t) &= \int_0^t v_{1,a,r}dt \\
V_{2,a,r}(t) &= \int_0^t v_{2,a,r}dt
\end{aligned}
\tag{1}
$$

where $N_{a,r}$ is the size of the population in age-group $a$ and region $r$. Parameters for vaccine waning $\omega_1 = 100^{-1}$ per day, $\omega_2 = 320^{-1}$ per day, $\Omega_1 = 360^{-1}$ per day and $\Omega_2 = 1500^{-1}$ per day are chosen to fit the recorded changes in efficacy over time (see Figure 3 of main Methods).

#### 2 Infection Dynamics

Within this section we detail the infection dynamics, including multiple exposed classes to generate an appropriate distribution for the generation time, the status of individuals with respect to their household, as well as variant and age-structure.

One of the key characteristics of the COVID-19 pandemic in the UK has been the use of self-isolation and household quarantining to reduce transmission. We approximate this process by distinguishing between first infections (caused by infection related to any non-household mixing) and subsequent household infections (caused by infection due to household mixing). We note that first infection really applies to any new infection brought into an infection free household. The first symptomatic case within a household has a probability ($H_t$) of leading to household quarantining; this curtails the non-household mixing of the individual and all subsequent infections generated by this individual. We use superscripts to denote the status of an infection with respect to this household structure: superscript $F$ refers to the first infection in a household that has not been quarantined; $SI$ and $SA$ refer to subsequent infections that are generated by a first infection that is symptomatic or asymptomatic respectively, again in a household that has not been quarantined; $QF$ refers to the first detected case in the household that leads to quarantining and $QS$ is all of their subsequent household infections.

We then use subscripts to denote the multiple stages withing the exposed class, the age-group $a$ of the infected individual, the region $r$ and the variant $\theta$:

$$
\begin{aligned}
\frac{dE^F_{1,a,r,\theta}}{dt} &= \lambda^F_{a,r,\theta}\mathcal{S}_{a,r} - 3\alpha_{r,\theta}E^F_{1,a,r,\theta} \\
\frac{dE^{SI}_{1,a,r,\theta}}{dt} &= \lambda^{SI}_{a,r,\theta}\mathcal{S}_{a,r} - 3\alpha_{r,\theta}E^{SI}_{1,a,r,\theta} \\
\frac{dE^{SA}_{1,a,r,\theta}}{dt} &= \lambda^{SA}_{a,r,\theta}\mathcal{S}_{a,r} - 3\alpha_{r,\theta}E^{SA}_{1,a,r,\theta} \\
\frac{dE^Q_{1,a,r,\theta}}{dt} &= \lambda^Q_{a,r,\theta}\mathcal{S}_{a,r} - 3\alpha E^Q_{1,a,r,\theta} \\
\frac{dE^X_{2,a,r,\theta}}{dt} &= 3\alpha_{r,\theta}E^X_{1,a,r,\theta} - 3\alpha_{r,\theta}E^X_{2,a,r,\theta} \\
\frac{dE^X_{3,a,r,\theta}}{dt} &= 3\alpha_{r,\theta}E^X_{2,a,r,\theta} - 3\alpha_{r,\theta}E^X_{3,a,r,\theta} \qquad (2)\\
\frac{dI^F_{a,r,\theta}}{dt} &= 3\bar{d}_a(1-H_t)\alpha_{r,\theta}E^F_{3,a,r,\theta} - \gamma_\theta I^F_{a,r,\theta} \\
\frac{dI^{SI}_{a,r,\theta}}{dt} &= 3\bar{d}_a\alpha_{r,\theta}E^{SI}_{3,a,r,\theta} - \gamma_\theta I^{SI}_{a,r,\theta} \\
\frac{dI^{SA}_{a,r,\theta}}{dt} &= 3\bar{d}_a(1-H_t)\alpha_{r,\theta}E^{SA}_{3,a,r,\theta} - \gamma_\theta I^{SA}_{a,r,\theta} \\
\frac{dI^{QF}_{a,r,\theta}}{dt} &= 3\bar{d}_a H_t\alpha_{r,\theta}E^F_{3,a,r,\theta} - \gamma_\theta I^{QF}_{a,r,\theta} \\
\frac{dI^{QS}_{a,r,\theta}}{dt} &= 3\bar{d}_a\alpha_{r,\theta}E^Q_{3,a,r,\theta} + 3d_{a,\theta}H_t\alpha_{r,\theta}E^{SA}_{3,a,r,\theta} - \gamma_\theta I^{QS}_{a,r,\theta} \\
\frac{dA^X_{a,r,\theta}}{dt} &= 3(1-\bar{d}_a)\alpha_{r,\theta}E^X_{3,a,r,\theta} - \gamma_\theta A^X_{a,r,\theta} \\
\frac{dR_{a,r}}{dt} &= \gamma_\theta\sum_{X,\theta}(I^X_{a,r,\theta} + A^X_{a,r,\theta}) \\
&\quad \text{where } X \in \{F, SI, SA, QF, QS\}
\end{aligned}
$$

$$
\begin{aligned}
\text{where } \lambda^F_{a,r,\theta} &= \sigma_a\widehat{\beta}_\theta\sum_b(\beta^S_{b,a,t} + \beta^W_{b,a,t} + \beta^O_{b,a,t})(I^F_{b,r,\theta} + I^{SI}_{b,r,\theta} + I^{SA}_{b,r,\theta} + \tau A^F_{b,r,\theta} + \tau A^{SI}_{b,r,\theta} + \tau A^{SA}_{b,r,\theta}) \\
\lambda^{SI}_{a,r,\theta} &= \sigma_a\widehat{\beta}_\theta\sum_b \beta^H_{b,a,t}I^F_{b,r,\theta}, \\
\lambda^{SA}_{a,r,\theta} &= \sigma_a\widehat{\beta}_\theta\sum_b \beta^H_{b,a,t}\tau_b A^F_{b,r,\theta}, \\
\lambda^Q_{a,r,\theta} &= \sigma_a\widehat{\beta}_\theta\sum_b \beta^H_{b,a,t}I^{QF}_{b,r,\theta}
\end{aligned}
$$

Here $\mathcal{S}$ is a measure of the susceptible population (including both naive, vaccinated and waned individuals); $\lambda$ refers to the force of infection generating first infections (superscript $F$), secondary infections within the home from a symptomatic or asymptomatic first infection (superscript $SI$ or $SA$) or from quarantined individuals (superscript $Q$); $\alpha$ is the rate of movement from exposed to infectious and $\gamma$ is the recovery rate; $\bar{d}$ is the probability of that an infection will be symptomatic (which is dependent on the vaccine status of infected individuals), and $H$ is the probably that a symptomatic infection will lead to household quarantining. The dependence of these parameters on age ($a$), region ($r$) and variant ($\theta$) is explicitly shown; only $H$ is time-dependent and is assumed to be related to the level of precautionary behaviour $\phi_t$.

The force of infection, $\lambda$, is again partitioned by whether the individual getting infected is the first, subsequent or from a quarantined household. This risk of infection is driven by the age-depending mixing matrices for home, school, work and other contacts ($\boldsymbol{\beta}^H$, $\boldsymbol{\beta}^S$, $\boldsymbol{\beta}^W$ and $\boldsymbol{\beta}^O$ respectively) which scale with the estimated time-dependent precautionary behaviour. The risk of infection also varies with the variant (as captured by $\widehat{\beta}_\theta$) and the age-dependent risk of infection ($\sigma_a$).

#### 2.1 Generic behaviour of the household quarantine model

Using this formation (equation 2) it has been shown that quarantining is able to reduce the reproductive ratio, $R$, below one even when there is strong within household transmission, as infection from quarantined individuals cannot escape the household [1]. Given the novelty of this additional household structure, we now clarify in more detail the action of this formulation. We give a simpler set of equations (based on a standard $SIR$ model) that contains a similar household structure; in particular, we take the standard $SIR$ model and split the infected class into those first infected within a household ($I_F$) and subsequent infections ($I_S$):

$$\begin{aligned}
\frac{dS}{dt} &= -[\beta^H I_F + \beta^O S(I_F + qI_S)]S \\
\frac{dI_F}{dt} &= \beta^O(I_F + I_S)S - \gamma I_F \\
\frac{dI_S}{dt} &= \beta^H I_F S - \gamma I_S \\
\frac{dR}{dt} &= \gamma(I_F + I_S)
\end{aligned}$$

Again, the transmission rate is split into within household transmission $\beta^H$ and all other transmission $\beta^O$ (i.e out-of-household transmission, which includes transmission at work or school). As in the full equations we assume that all within-household infection is generated by the first case, which allows us to greatly simplify the unfolding dynamics. Here for even greater simplicity we ignore the action of household quarantining (by setting $q = 1$) and instead focus on the dynamics as $\beta^O$ is reduced. We compare this new formulation to the standard SIR model without this additional structure:

$$\begin{aligned}
\frac{dS}{dt} &= -\widehat{\beta}^H IS - \widehat{\beta}^O IS \\
\frac{dI}{dt} &= \widehat{\beta}^H IS + \widehat{\beta}^O IS - \gamma I \\
\frac{dR}{dt} &= \gamma I
\end{aligned}$$

where we retain the split in transmission type for ease of comparison.

The early growth rate of the two models are $\widehat{r} = \widehat{\beta}^H + \widehat{\beta}^O - \gamma$ for the simple SIR model, and $r = \frac{1}{2}\left[\beta^O - 2\gamma + \sqrt{\beta^{O^2} + 4\beta^O\beta^H}\right]$ for the household structured version. From this simple comparison, it is clear that for the simple model the growth rate can remain positive even when control measures substantially reduce transmission outside the home ($\widehat{\beta}^O$ gets reduced); the growth rate will remain positive so long as $\widehat{\beta}^H > r$. In contrast for the structured version there is always a threshold level of transmission outside the household ($\beta^O_c = \gamma^2/(\beta^H + \gamma)$) that is needed to maintain positive growth. As such the structured model is better able to capture the actions of quarantining and isolation, reducing the growth rate even when there is substantial within-household transmission.

For both the simple model given here and the full COVID-19 model, the inclusion of this additional household structure reduces the amount of within-household transmission compared to a model without this structure — as only the initial infection in each household ($I_F$) generates secondary within-household cases. It is therefore necessary to rescale the household transmission rate $\beta^H$, compared to the simple model, to obtain the appropriate average within-household attack rate. For the full COVID-19 model, we find that a simple multiplicative scaling to the household transmission ($\beta^H \approx z\widehat{\beta}^H$, with $z = 1.3$) generates a comparable match between the new model and a model without this household structure – even when age structure is included.

### 3 Linking Infection, Vaccination Dynamics and Outcomes

To link together the previous two model sections, we need to focus on the status of those individuals who get infected. We define a vector of potentially susceptible groups:

$$\mathbf{\Psi}_{a,r} = (S_{a,r}, VS_{1,a,r}, VS_{2,a,r}, WS_{1,a,r}, WS_{2,a,r}, R_{a,r}, WR_{1,a,r}, WR_{2,a,r})$$

The total susceptible variable that feeds into the infection equations (2) is then the dot product of this susceptible vector with the vector of susceptibility to infection:

$$\mathcal{S}_{a,r} = \mathbf{\Psi}_{a,r} \cdot \left(1, s^I_1, s^I_2, s^I_2, s^I_W, 0, 0, s^I_R\right)$$

noting that the susceptible in the first waned compartment ($WS_1$) is the same as for those who have received both doses of vaccine, and that the risk of infection to those in the recovered or first waned compartment after recovery ($WR_1$) is zero. The values of the susceptibility $s^I$ is one minus the protection against infection (as given in Table 2 of the Methods section), with the weighting between the protection afforded by AstraZeneca and by mRNA vaccines (Pfizer or Moderna) given by the age and region specific ratio of the vaccines delivered up to that point in time.
The risk of displaying symptoms ($\overline{d}_a$) used in equation (2) is also based on the vector of susceptiblities:

$$\overline{d}_a = D_a \ \mathbf{\Psi}_{a,r} \cdot \left(1, s^d_1, s^d_2, s^d_2, s^d_W, 0, 0, s^d_R\right)$$

where $s^d$ is the ratio of one minus the protection against symptoms relative to the one minus the protection against infection, and $D_a$ is the age-dependent risk of developing symptoms (extracted from the early case-reporting data [1]).

A similar approach is used to determine the number of hospital admission, the number of ICU admissions, the number of deaths and the level of hospital and ICU occupancy. We first define the rate of generating newly symptomatic infectious individuals as:

$$NI_{a,r,\theta}(t) = 3\overline{d}_a \alpha_{r,\theta} \sum_X E^X_{3,a,r,\theta}(t)$$

The modelled rate of admission to hospital or admission to ICU is then given by:

$$\begin{aligned} M^{HospAd}_{a,r,\theta}(t) &= \int_0^\infty H_a \widehat{H}_{r,\theta} T^H(\tau) NI_{a,r,\theta}(t-\tau) d\tau \\ M^{ICUAd}_{a,r,\theta}(t) &= \int_0^\infty I_a \widehat{I}_{r,\theta} T^I(\tau) NI_{a,r,\theta}(t-\tau) d\tau \end{aligned}$$

where each term consists of an age-dependent risk ($H_a$ and $I_a$, taken from the early data), a regional and variant dependent scaling factor ($\hat{H}_{r,\theta}$ and $\hat{I}_{r,\theta}$, estimated through our fitting procedures) and a delay between the onset of symptoms and the epidemiological event ($T^H(\tau)$ and $T^I(\tau)$, with these distributions based on recorded data). The modelled death rate follows a similar form, but is amplified by high levels of hospital occupancy in a region relative to the population size ($M_r^{HospOcc}/N_r$):

$$M_{a,r,\theta}^{Death}(t) = \left(1 + F_r \frac{M_r^{HospOcc}}{N_r}\right) \int_0^\infty D_a \widehat{D}_{r,\theta} T^D(\tau) NI_{a,r,\theta}(t-\tau) d\tau$$

where $F_r$ is estimated at 1600 (CI 770-2800), such that the death rate approximately doubles if 0.06% of a region is in hospital - which is close to the levels observed at the peak of the Alpha wave.

Hospital and ICU occupancy are then computed based on the recorded distributions ($D$) of length of stays [1]:

$$\begin{aligned} M_{a,r,\theta}^{HospOcc}(t) &= \int_0^\infty D_a^H(\tau\kappa_{r,\theta}^H) M_{a,r,\theta}^{HospAd}(t-\tau) d\tau \\ M_{a,r,\theta}^{ICUOcc}(t) &= \int_0^\infty D_a^I(\tau\kappa_{r,\theta}^I) M_{a,r,\theta}^{ICUAd}(t-\tau) d\tau \end{aligned}$$

These distributions are scaled for each age-group and variant (by a factor $\kappa$); We note that children and young adults spending less average time in hospital and the average length of stay being longer for Alpha and Delta variants than for the wildtype variant.

#### 4 Precautionary Behaviour

One of the main parameters that drives much of the dynamics is the level of precautionary behaviour $\phi_{t,r}$. We view $\phi_{t,r}$ as slowly varying (expect when there is an abrupt change in policy) and captures how the risky interactions between susceptible and infectious individuals scale throughout the pandemic. As such the precautionary behaviour captures the changes in social mixing (including working from home) as well as behavioural changes such as mask-use and isolation. The level of precautionary behaviour is estimated for each week and each region as part of our fitting procedure (see below) and is a scalar parameter between zero and one; when $\phi = 0$ we have returned to pre-pandemic mixing whereas $\phi = 1$ corresponds to a stringent lock-down. In particular, $\phi_t$ is used to regulate the home, work, school and other transmission matrices:

$$\begin{aligned} \beta_{b,a,t}^H &= \tilde{\beta}_{a,b}^H \left[(1-\phi_t) + \phi_t q^H\right] \\ \beta_{b,a,t}^S &= \tilde{\beta}_{a,b}^S \left[(1-\phi_t) + \phi_t q^S\right] \\ \beta_{b,a,t}^W &= (1-f)\tilde{\beta}_{a,b}^W \left[(1-\phi_t) + \phi_t q^W\right] + f\tilde{\beta}_{a,b}^W \left((1-\phi_t) + \phi_t q^W\right)\left((1-\phi_t) + \phi_t q^O\right) \\ \beta_{ba}^O &= \tilde{\beta}_{b,a}^O ((1-\phi_t) + \phi_t q^O)^2 \end{aligned}$$

Here, $\tilde{\beta}$ are the mixing matrices for home, school, work and other contacts during pre-pandemic circumstances as given by [2]; and $q$ acts to define the scaling during severe retardation of social mixing ($q^H = 1.25$ such that within household mixing increases during lockdown, $q^W = 0.2$ such that some work activities have to continue, $q^S = 0.05$ and $q^O = 0.05$). For work contacts we separate industries that are public-facing ($f = 0.3$, such a leisure and retail) from other employment; contacts in public-facing industries are assumed to scale with quadratically accounting for both the number of individuals at work and the number of people accessing these activities.

While the level of precautionary behaviour is inferred by matching to epidemiological data, it is interesting to compare the estimates to other more directly recorded measures of behaviour. In Figure S1 we compare our estimated values to google-mobility data [3] (top two panels) and diary-based

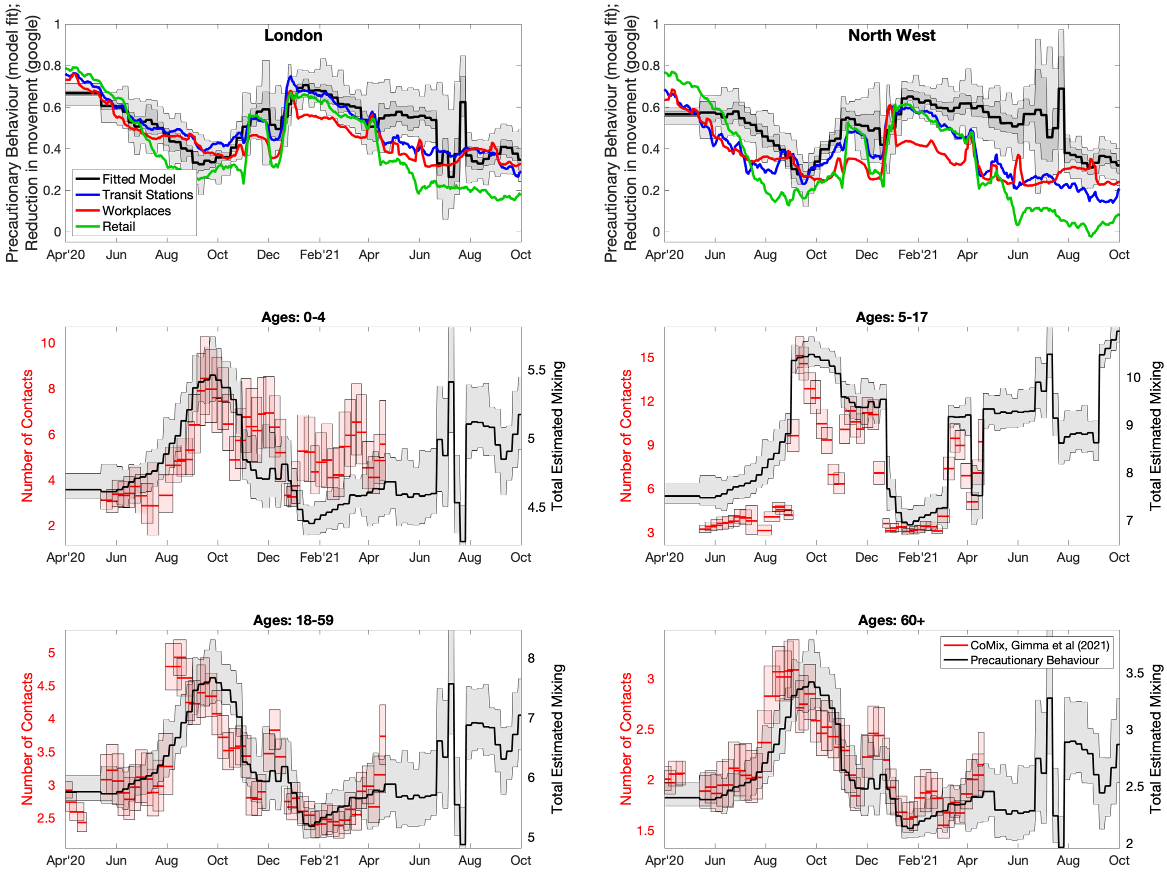


**Fig. S1: Comparison between estimated measures of social mixing with the model (black) and recorded observations from google mobility [3] and CoMix [4].** In the top two panels we directly compare the level of precautionary behaviour $\phi_{t,r}$ with the reduction in movements (transit stations in blue, workplaces in red and retail in green) as estimate from google mobility [3] observations. This is shown for London and the North West; there is better qualitative agreement for London which may reflect the degree to which recorded movements capture population-level mixing in the capital. In the lower four panels we qualitatively compare the number of contacts recorded by CoMix [4] in four age-groups (0-4, 5-17, 18-59 and 60+) with the estimated mixing from our age-structured transmission matrices. Given the mixing matrices are re-scaled to generated a transmission rate we would not expect a one-to-one agreement with the recorded number of contacts.

records of contacts [4] (lower four panels). For the google mobility comparison, we consider the estimated precautionary behaviour ($\phi_t$) in comparison to the reduction in movements as measures by google over this period. We focus on two regions, London and the North West, noting that while both have similar features, the qualitative agreement for London is far stronger. When comparing to the Co-Mix study [4], the available data is number of contacts per person in a given age-group, we therefore compare this to the mixing matrices for those ages (e.g. $\sum_b \beta^H_{b,a,t} + \beta^S_{b,a,t} + \beta^W_{b,a,t} + \beta^O_{b,a,t}$ for age group $a$), noting that as these measure different things there will not be a one-to-one matching. We again find that our estimated mixing agrees with the age-structured trends identified by Co-Mix. A more thorough description of the Precautionary Behaviour and the different behavioural elements is given in [5].

### 5 Parameter Inference

Key to the accuracy of any model are the parameters that underpin the dynamics. With a model of this complexity, a large number of parameters are required. Some, such as vaccine efficacy and waning, are taken from current literature; while others are inferred from the epidemic dynamics.

Of the inferred parameters there are three basic classes; those, such as scalings of the case-hospitalisation ratios, that are different between regions and variants; others such as age-dependent susceptiblity are universal (the same for all regions and variants); while the level of precautionary behaviour over time changes on a weekly time-scale. Bayesian inference, using an MCMC process, is applied to each of the seven NHS regions in England to determine posterior distributions for each of the regional parameters (further details are given in [6]). The distribution of parameters leads to uncertainty in model projections, which is represented by the 95% prediction interval in all graphs (this interval contains 95% of all predictions). We note that when we compare two scenarios (for example vaccination with a 3-week interval, with vaccination using a 12-week interval) we compared simulations with the same parameters chosen from the posterior distributions - and then calculated means and 95% prediction intervals based on these results.

As the epidemic has progressed, new posterior distributions based on the latest data are initialised from previous MCMC chains – ensuring a rapid fit to historical data. In general this refitting process has been performed weekly (or twice weekly) throughout the pandemic. For the time period of relevance in this paper (December 2020 - September 2021), we matched to seven observations: hospital admissions, hospital occupancy, ICU occupancy (noting that data on ICU admissions is not available), deaths, proportion of pillar 2 (community) test that are positive, the proportion of pillar 2 tests that are S-gene negative (as a signal of the ratio of wild-type to Alpha variant, then a signal of the ratio of Delta to Alpha variant), and the early REACT data as a measure of sero-prevalence [7]. As such our log-likelihood function is given by:

$$
\begin{aligned}
\text{LogLike}(\text{Data}_r|\boldsymbol{\Theta}_r) \;=\; & \prod_t lP(\text{Hospital Admissions}_r(t)|\sum_{a,\theta} M^{HospAd}_{a,r,\theta}(t)) \;+ \\
& lP(\text{Deaths}_r(t)|\sum_{a,\theta} M^{Death}_{a,r,\theta}(t)) \;+ \\
& lP(\text{Hospital Occupancy}_r(t)|\sum_{a,\theta} M^{HospOcc}_{a,r,\theta}(t)) \;+ \\
& lP(\text{ICU Occupancy}_r(t)|\sum_{a,\theta} M^{ICUOcc}_{a,r,\theta}(t)) \;+ \\
& lB(\text{Positive Tests}_r(t)|\text{Total Tests}_r(t), \sum_{a,\theta} F_\theta NI_{a,r,\theta}(t)/\sum_a N_{a,r}) \;+ \\
& lB(\text{S-gene Neg Tests}_r(t)|\text{Total Tests}_r(t), \sum_a NI_{a,r,Alpha}(t)/\sum_{a,\theta} NI_{a,r,\theta}(t)) \;+ \\
& lB(\text{REACT Sero}_r(t)|\text{REACT Tests}_r(t), \sum_{a,\theta}\int_0^t NI_{a,r,\theta}(\tau)d\tau/\sum_a N_{a,r})
\end{aligned}
$$

where $lP$ and $lB$ are the logs of Poisson and Binomial probabilities, $N_{a,r}$ is the population size of individuals in age-group $a$ and region $r$, $F_\theta$ is a variant dependent level of reporting, and $\boldsymbol{\Theta}$ is the set of all model parameters for a given region. We note that in [6], which was written in the early stages of the pandemic, we did not fit to S-gene data as we had been dealing with a single variant. Although not part of the underlying transmission dynamics, the seven quantities for each spatial region can be generated from the number, age and type of infection within the model, as described above.

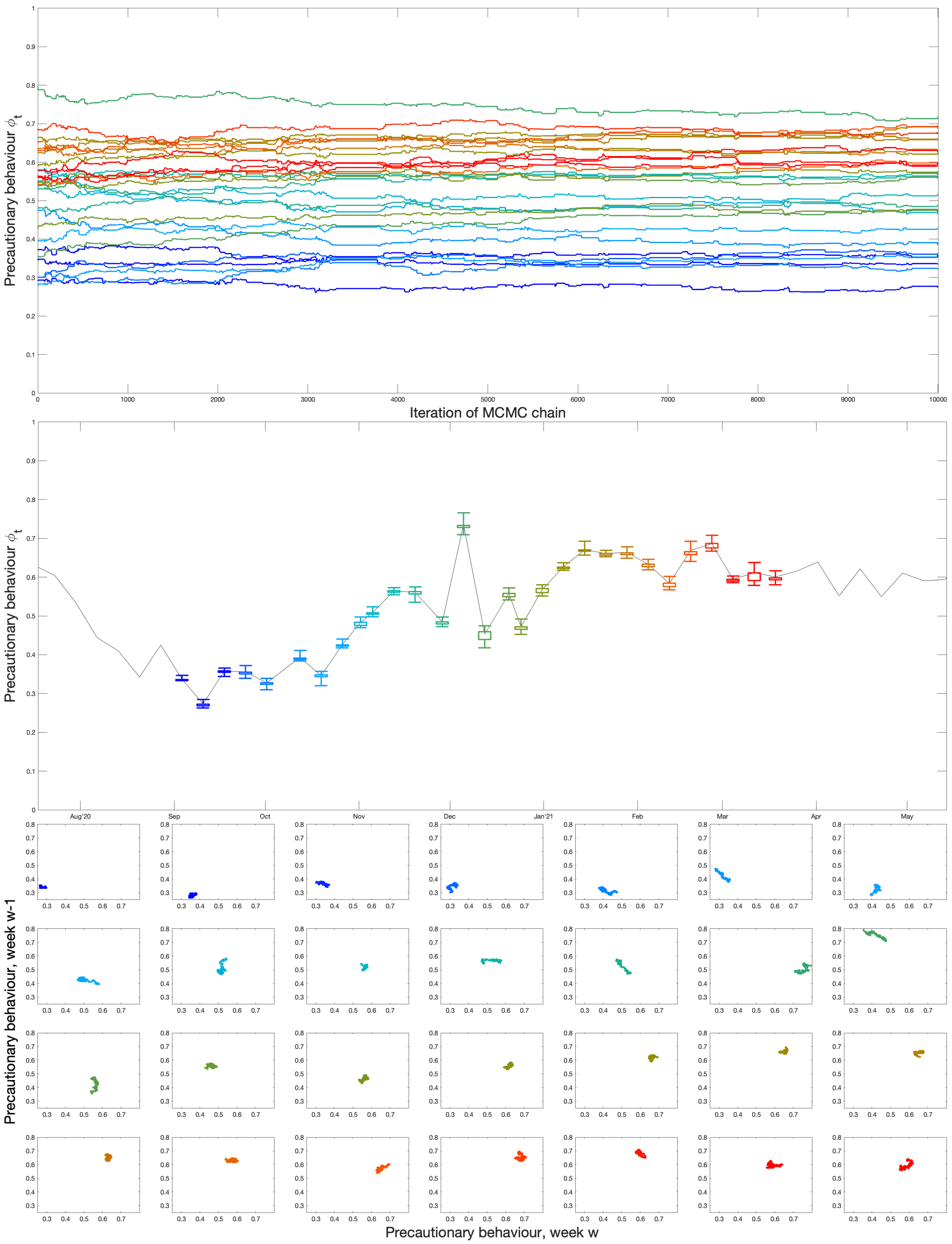


**Fig. S2: Example of an MCMC chain for $\phi_t$ in the Midlands from the main Alpha wave.** Throughout, the plotted data is colour-coded (from blue to red) corresponding to the date it represents. The top panel show the behaviour of the 29 weekly values of $\phi$ across 10,000 iterations of the chain; the central panel shows the mean $\phi$ values (line) together with the inter-quartile and 95% credible interval for each week; while the lower panel shows the correlation between the value of $\phi$ against the value the previous week.

Figure S2 shows an example of one chain for the precautionary behaviour, $\phi_{t,r}$, during the period September 2020 to March 2021 that captures the bulk of the Alpha wave. The chain is 10,000 iterations of the MCMC process, and refers to the precautionary behaviour for the Midlands region; a typical chain from the weekly re-fitting is around 3-5,000 iterations. The figure shows the individual chains for the 29 weekly values of $\phi_t$ (top panel); the mean, inter-quartile range and 95% credible intervals for each parameter through time; and the correlation between $\phi_t$ and the value the preceding week.

### 6 Peak Demand

In the main paper we have focused on the total number of hospital admissions and deaths as an appropriate public health measure, with the applied aim of minimising both. However, the burden on health services is often more closely related to the peak numbers. Here we compare the peak numbers of deaths, hospital admissions and hospital occupancy, reporting values for the Alpha (December 2020 to May 2021) and Delta (May to September 2021) waves within our simulation intervals.

**Table S1:** Peak in deaths, hospital admissions and hospital occupancy in England in two periods corresponding to the Alpha wave (from 8th December 2020 to 15th May 2021) and Delta waves (from 16th May 2021 to 1st September 2021), as observed and from four model scenarios as described in the main text. For each model scenario we give the mean and 95% prediction intervals from 400 samples of the posterior distribution. It should be noted that all model results assume the same relaxation of control measures throughout 2021.

| **Alpha, December 2020 - May 2021** | | | |
|---|---|---|---|
| | Deaths | Hospital Admissions | Hospital Occupancy |
| Observed | 1250 | 3818 | 31459 |
| (i) Default | 1170 (1120-1230) | 3710 (3610-3810) | 31,500 (30900-31900) |
| (ii) Prioritise youngest | 1270 (1220-1340) | 3900 (3790-4000) | 33,100 (32,500-33,500) |
| (iii) 3-week interval, default efficacy | 1170 (1120-1230) | 3710 (3610-3810) | 31,500 (30,900-31,900) |
| (iv) 3-week interval, lower efficacy | 1170 (1120-1230) | 3710 (3610-3820) | 31,500 (30,900-31,900) |
| **Delta, May 2021 - September 2021** | | | |
| | Deaths | Hospital Admissions | Hospital Occupancy |
| Observed | 108 | 851 | 6057 |
| (i) Default | 120 (110-140) | 820 (790-870) | 6100 (5900-6300) |
| (ii) Prioritise youngest | 220 (10-2040) | 570 (20-4670) | 4200 (400-34,400) |
| (iii) 3-week interval, default efficacy | 220 (180-260) | 1800 (1580-2080) | 11,200 (9800-12,800) |
| (iv) 3-week interval, lower efficacy | 340 (250-450) | 2360 (1990-2980) | 14,700 (12,200-18,400) |

As expected from the results of the main paper, changing to a three-week interval does not substantially change the Alpha variant peak (which occurred shortly after vaccination had begun), but does lead to an increase in the peaks for all three measures during the Delta waves. In contrast, vaccinating the youngest first leads to a slightly larger Alpha variant peak and a larger peak in deaths during the Delta wave; the mean hospital admission and hospital occupancy peaks are reduced, although the upper 95% prediction interval is larger due to greater variability between replicates.

### 7 Comparison between Model and Data for Age-structured Severe Disease

It has been clear throughout the pandemic that age-structure plays a major role in both the acquisition of infection and the severity of disease - with the risk of hospital admission and death increasing rapidly with age. In the main text we compare model results with data for the entire population, focusing on the total daily number of hospital admissions (associated with a positive COVID-19 test) and the total number of daily deaths (within 28 days of a positive test). Here we extend this comparison, plotting data and model results on the same figure for six different age ranges (Fig. S3 and Fig. S4). We note that the age-groups for which hospital admissions are reported do not necessarily correspond with the 5-year age bins used within the model simulations. We assume homogeneity within an age-group in the model (i.e. all individual aged 0 to 4 have the same mean risk of infection and hospital admission); this means that when computing the expected number of daily admissions for those aged 6-17, for example, we sum 80% of the second age-group in the model (age 5-9), 100% of the third age-group (age 10-14) and 60% of the forth age-group (age 15-19). For deaths, from the empirical data we know the age of each person in years and so are able to make a comparison to simulated outputs using an amalgamation of 5-year age groups.

It should be noted, as described above, that the inference processes only use aggregate (non-age-structured) data to determine the match between model and the unfolding epidemic. The exception is the risk of hospital admission and death following infection, which for each variant of concern are matched to the total reported for each age group. In general, we find that during the second wave (December 2020 - March 2021) the model tends to slightly under-estimate hospital admissions and over-estimate deaths in the 45-64 age group. However, it captures the bulk properties of the waves, especially the dominance of severe disease in older age-groups.

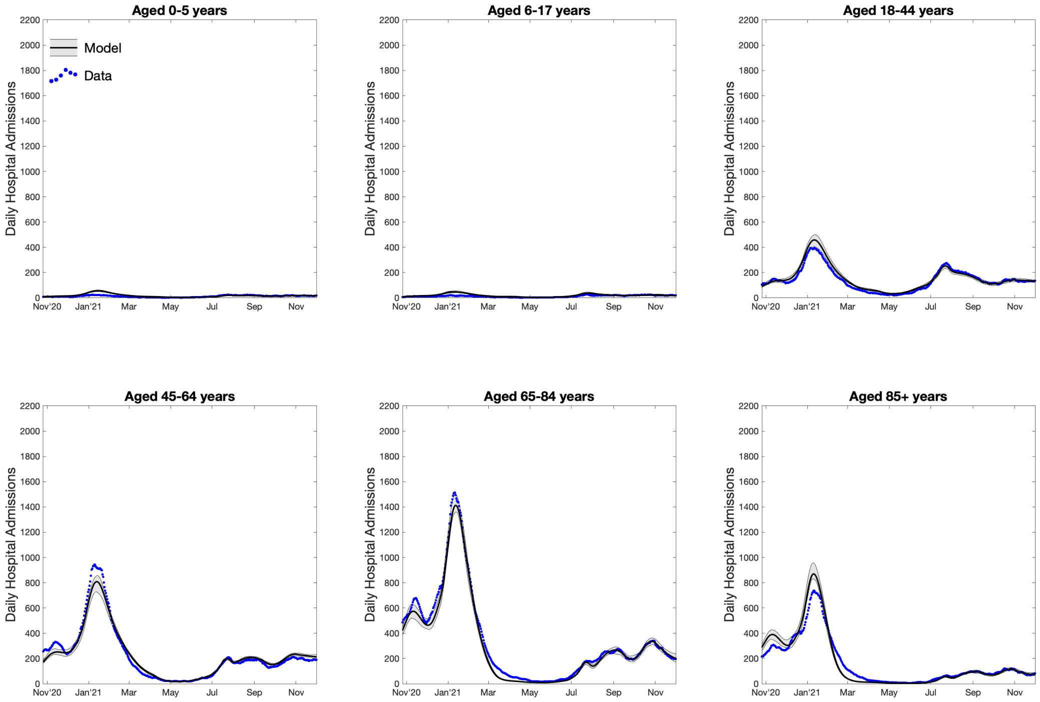


**Fig. S3: Comparison between models and data for age-structured hospital admissions.** We depict the data as solid black dots and results from the model simulations in blue - no attempt is made to add the appropriate noise to these raw deterministic simulations. The solid lines corresponds to mean values and the shaded area shows the 95% prediction interval (i.e. it contains 95% of all predictions at each point in time).

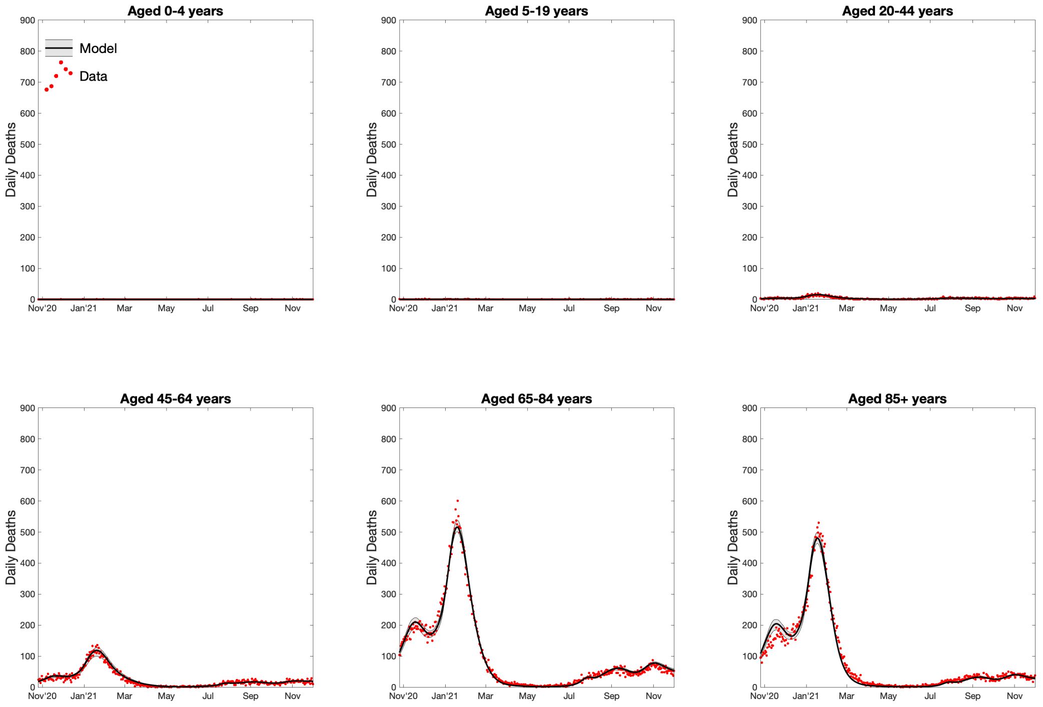


**Fig. S4: Comparison between models and data for age-structured deaths.** We depict the data as solid black dots and results from the model simulations in red- no attempt is made to add the appropriate noise to these raw deterministic simulations. The solid lines corresponds to mean values and the shaded area shows the 95% prediction interval (i.e. it contains 95% of all predictions at each point in time).

### 8 Comparison between Model and Data for the Seven NHS Regions of England

A similar comparison can be made against the model and data for the seven NHS regions (and England as a whole).

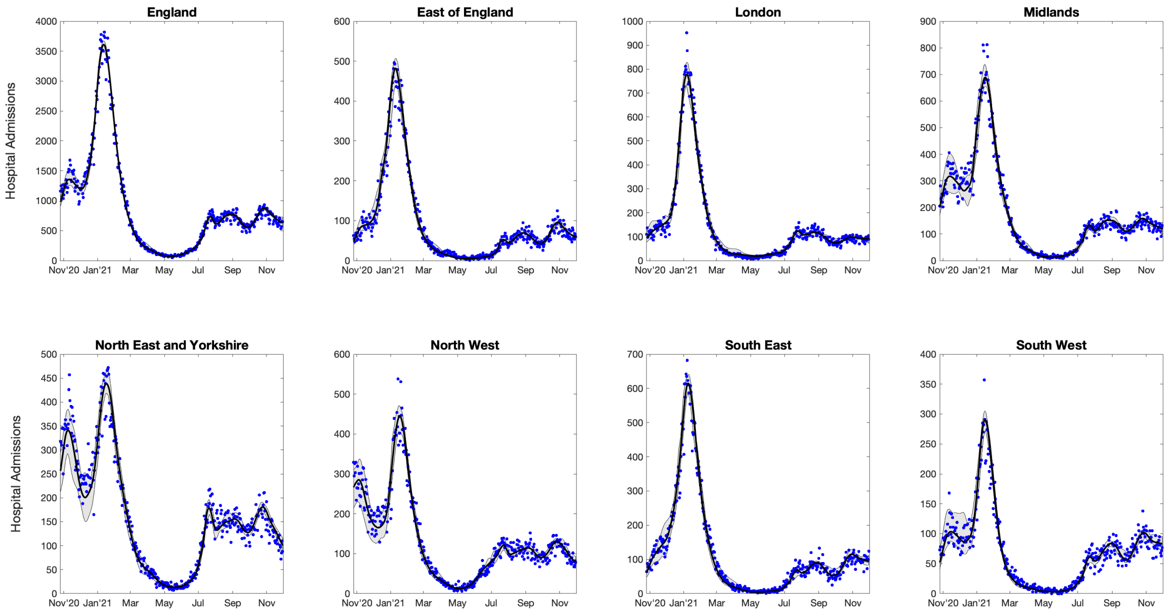


**Fig. S5: Comparison between models and data for regional hospital admissions.** We depict the data as solid black dots and results from the model simulations in blue - no attempt is made to add the appropriate noise to these raw deterministic simulations. The solid lines corresponds to mean values and the shaded area shows the 95% prediction interval (i.e. it contains 95% of all predictions at each point in time).

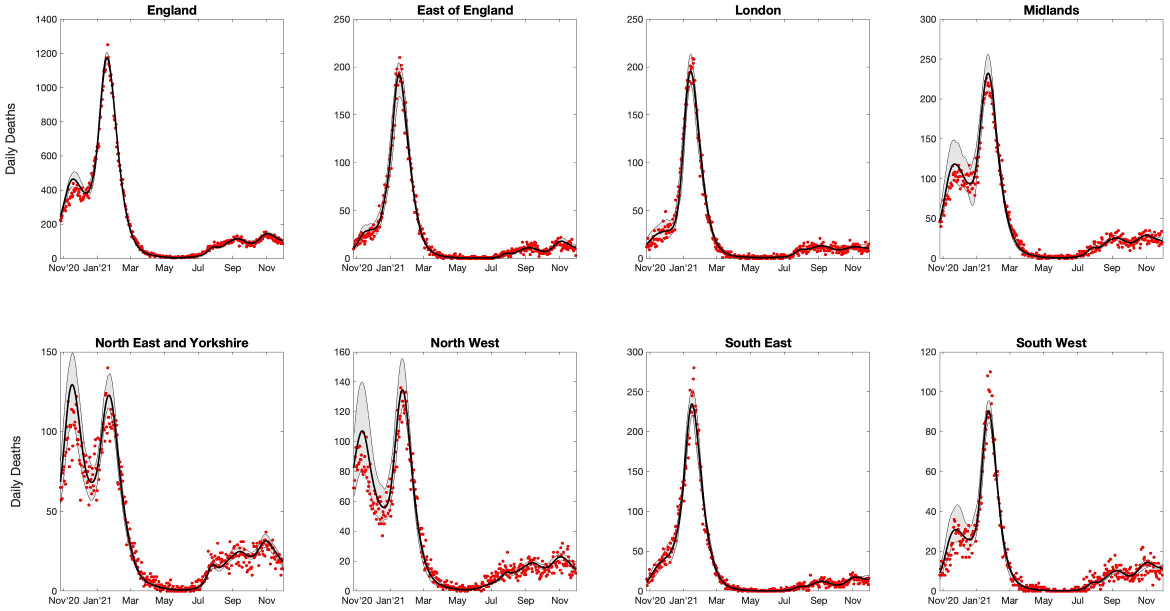


**Fig. S6: Comparison between models and data for regional deaths.** We depict the data as solid black dots and results from the model simulations in red- no attempt is made to add the appropriate noise to these raw deterministic simulations. The solid lines corresponds to mean values and the shaded area shows the 95% prediction interval (i.e. it contains 95% of all predictions at each point in time).

#### 9 Sensitivity to Vaccine Efficacy Parameters

Most of the model parameters are inferred by matching model output to available regional data. The exception are the parameters governing vaccine efficacy, which generally require individual level linked datasets that are able to compare levels of infection, severe disease and mortality between vaccinated and unvaccinated individuals. Our efficacy values have been taken from a range of UKHSA documents produced over the course of the pandemic [8]. Here we consider sensitivity to these vaccine efficacy input parameters, noting that each alternative set of vaccine efficacy values requires that the model is re-fit to the post-vaccination data. This refitting precludes an exhaustive sensitivity analysis. Instead, we consider the point estimates, lower and upper confidence intervals as given in the SAGE subcommittee consensus document [9]. (In this analysis we have not changed the level of onward transmission following infection in vaccinated individuals.) We assume efficacies either take the lower (or upper) confidence interval for all vaccine efficacy parameters, thereby generating extreme lower (or upper) bounds on the impact of vaccination. For each set of vaccine efficacies, we refit the model and generate the results shown in Fig. 1 of the main paper. We present the vaccine parameters used and the model results in the same format as Fig. 1, before providing a tabulated set of results that allows for a more direct comparison.

#### 9.1 Mid-Point Efficacy Estimates

**Table S2:** Vaccine efficacy parameters corresponding to midpoint estimates of the SAGE Vaccine Effectiveness Expert Panel subgroup [9]. We display the assumed percentage protection from the two vaccine types against the Alpha and Delta variants after one dose (D1), after two doses (D2, shown for the dose interval gaps of 12-weeks/3-weeks), and after waning (W). These values correspond to relatively slow waning of vaccine protection as in the main text ($\omega_1 = 100^{-1}$ per day, $\omega_2 = 320^{-1}$ per day, $\bar{\omega} = 420^{-1}$ per day) [10].

| Protection Against | AstraZeneca | | | | | | Pfizer/Moderna | | | | | |
|---|---|---|---|---|---|---|---|---|---|---|---|---|
| | Alpha | | | Delta | | | Alpha | | | Delta | | |
| | D1 | D2 | W | D1 | D2 | W | D1 | D2 | W | D1 | D2 | W |
| Infection | 60 | 80/65 | 0 | 40 | 65/48 | 0 | 60 | 85/77 | 0 | 55 | 90/80 | 0 |
| Symptoms | 60 | 80/66 | 0 | 45 | 65/49 | 0 | 60 | 90/82 | 0 | 55 | 90/85 | 0 |
| Hospital | 80 | 95 | 70 | 80 | 95 | 70 | 80 | 95 | 70 | 80 | 99 | 70 |
| Mortality | 80 | 95 | 70 | 80 | 95 | 70 | 80 | 95 | 70 | 80 | 99 | 70 |
| Transmission | 45 | 45 | 20 | 30 | 30 | 20 | 45 | 45 | 20 | 30 | 30 | 20 |

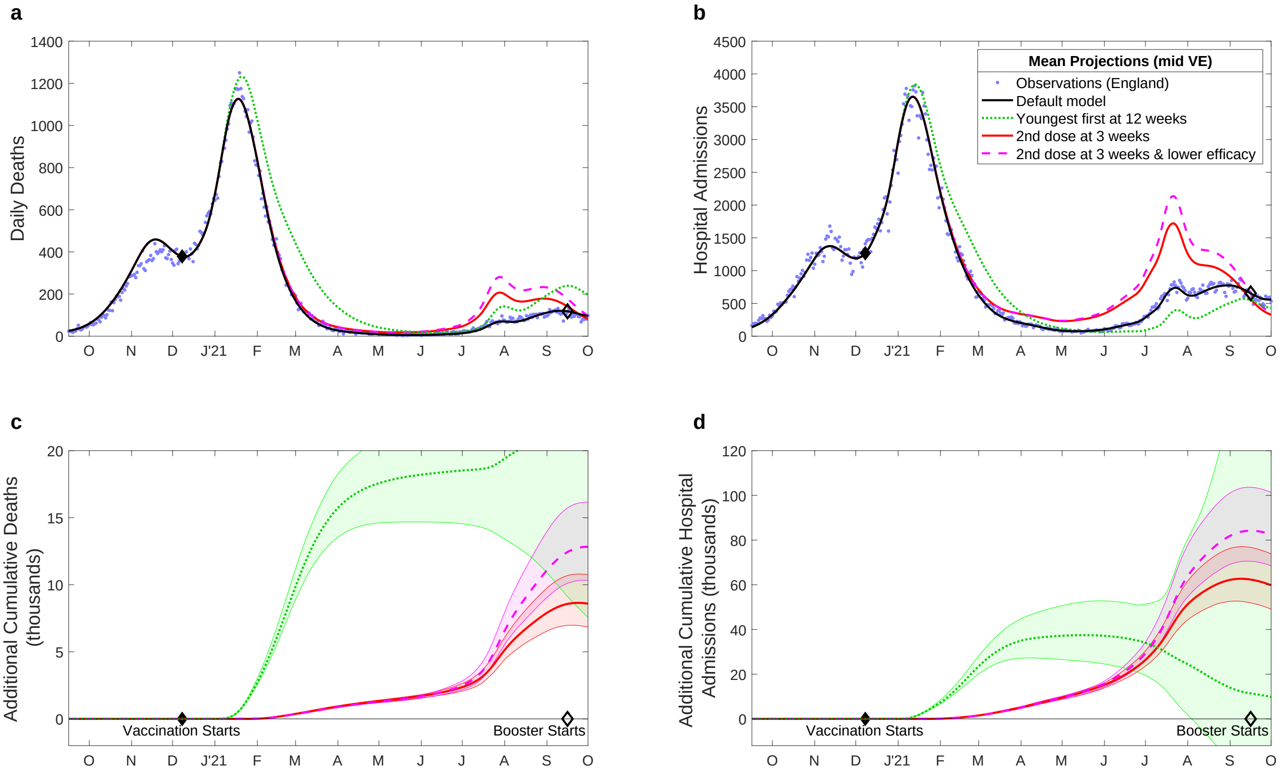


**Fig. S7:** Projected changes in daily deaths (**a**, **c**) and hospital admissions (**b**,**d**) with differing vaccination patterns. This is the equivalent of Fig. 1 in the main text, but uses the mid-point estimates of vaccine efficacy from the SAGE Vaccine Effectiveness Expert Panel subgroup [9].

#### 9.2 Lower Efficacy Estimates

**Table S3:** Vaccine efficacy parameters corresponding to the lower bounds of the SAGE Vaccine Effectiveness Expert Panel subgroup [9]. We display the assumed percentage protection from the two vaccine types against the Alpha and Delta variants after one dose (D1), after two doses (D2, shown for the dose interval gaps of 12-weeks/3-weeks), and after waning (W). These values correspond to relatively slow waning of vaccine protection as in the main text ($\omega_1 = 100^{-1}$ per day, $\omega_2 = 320^{-1}$ per day, $\bar{\omega} = 420^{-1}$ per day) [10].

| Protection Against | AstraZeneca | | | | | | Pfizer/Moderna | | | | | |
|---|---|---|---|---|---|---|---|---|---|---|---|---|
| | Alpha | | | Delta | | | Alpha | | | Delta | | |
| | D1 | D2 | W | D1 | D2 | W | D1 | D2 | W | D1 | D2 | W |
| Infection | 55 | 65/55 | 0 | 30 | 60/37 | 0 | 55 | 65/55 | 0 | 40 | 80/72 | 0 |
| Symptoms | 55 | 70/55 | 0 | 40 | 60/40 | 0 | 55 | 85/78 | 0 | 50 | 80/72 | 0 |
| Hospital | 75 | 80 | 70 | 75 | 90 | 70 | 75 | 90 | 70 | 75 | 90 | 70 |
| Mortality | 75 | 80 | 70 | 75 | 85 | 70 | 90 | 80 | 70 | 75 | 90 | 70 |
| Transmission | 45 | 45 | 20 | 30 | 30 | 20 | 45 | 45 | 20 | 30 | 30 | 20 |

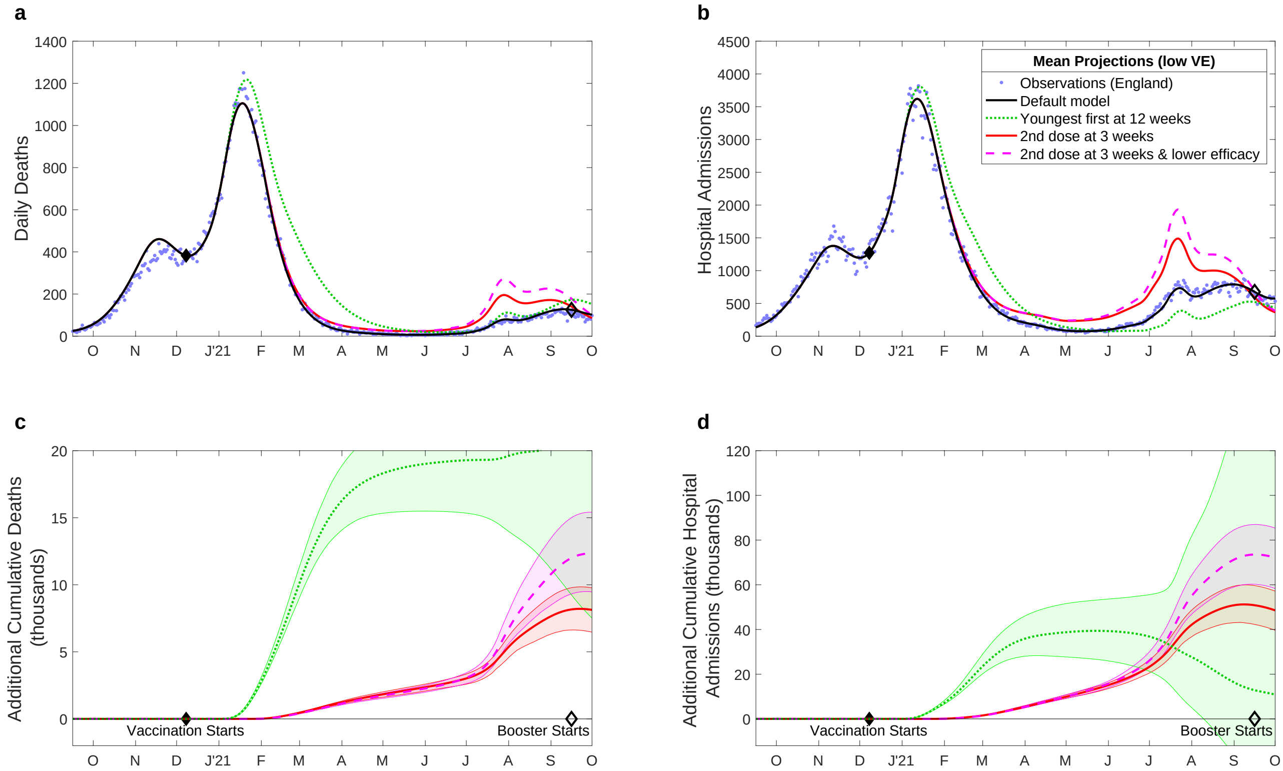


**Fig. S8:** Projected changes in daily deaths (**a**, **c**) and hospital admissions (**b**,**d**) with differing vaccination patterns. This is the equivalent of Fig. 1 in the main text, but uses the mid-point estimates of vaccine efficacy from the SAGE Vaccine Effectiveness Expert Panel subgroup [9].

#### 9.3 Upper Efficacy Estimates

**Table S4:** Vaccine efficacy parameters corresponding to the upper bounds of the SAGE Vaccine Effectiveness Expert Panel subgroup [9]. We display the assumed percentage protection from the two vaccine types against the Alpha and Delta variants after one dose (D1), after two doses (D2, shown for the dose interval gaps of 12-weeks/3-weeks), and after waning (W). These values correspond to relatively slow waning of vaccine protection as in the main text ($\omega_1 = 100^{-1}$ per day, $\omega_2 = 320^{-1}$ per day, $\bar{\omega} = 420^{-1}$ per day) [10].

| Protection Against | AstraZeneca | | | | | | Pfizer/Moderna | | | | | |
|---|---|---|---|---|---|---|---|---|---|---|---|---|
| | Alpha | | | Delta | | | Alpha | | | Delta | | |
| | D1 | D2 | W | D1 | D2 | W | D1 | D2 | W | D1 | D2 | W |
| Infection | 70 | 90/77 | 0 | 50 | 75/55 | 0 | 70 | 90/84 | 0 | 70 | 95/92 | 0 |
| Symptoms | 70 | 90/77 | 0 | 55 | 75/55 | 0 | 70 | 95/92 | 0 | 65 | 95/92 | 0 |
| Hospital | 85 | 99 | 70 | 85 | 99 | 70 | 85 | 99 | 70 | 85 | 99 | 70 |
| Mortality | 85 | 99 | 70 | 85 | 99 | 70 | 85 | 99 | 70 | 85 | 99 | 70 |
| Transmission | 45 | 45 | 20 | 30 | 30 | 20 | 45 | 45 | 20 | 30 | 30 | 20 |

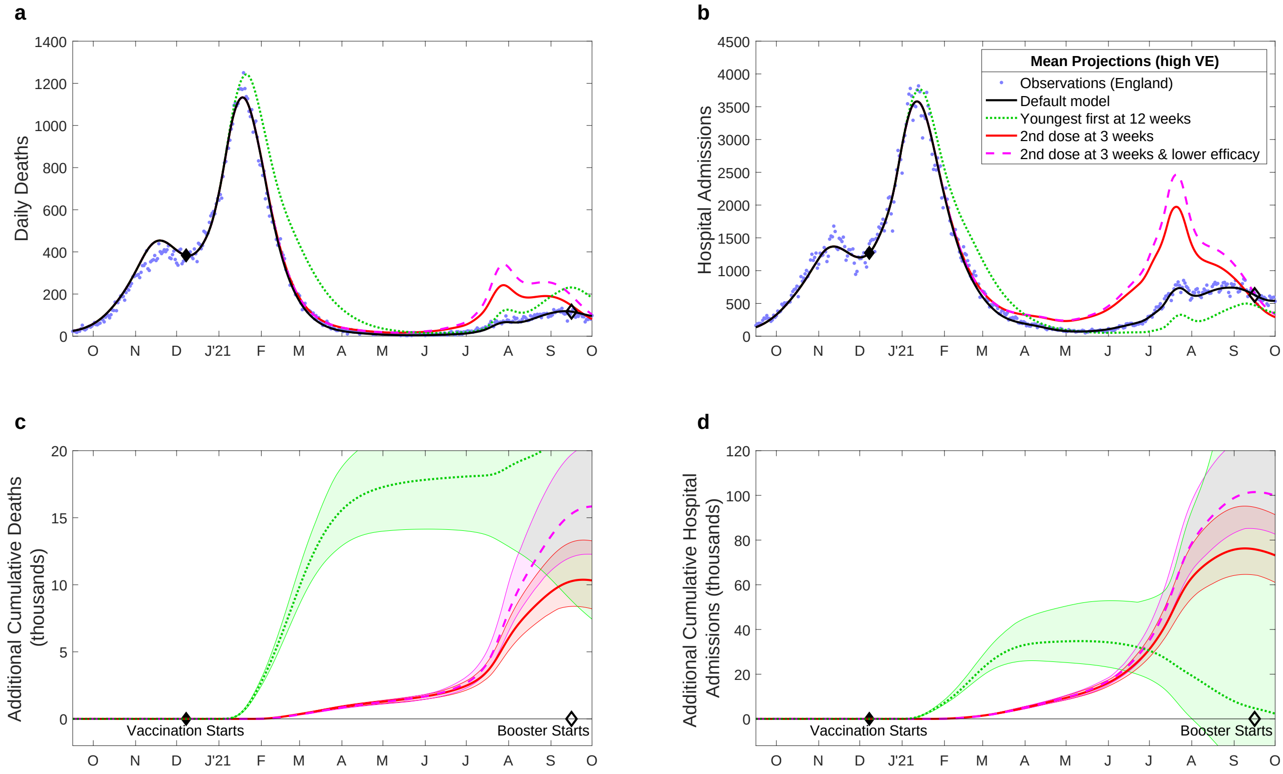


**Fig. S9:** Projected changes in daily deaths (**a**, **c**) and hospital admissions (**b**,**d**) with differing vaccination patterns. This is the equivalent of Fig. 1 in the main text, but uses the mid-point estimates of vaccine efficacy from the SAGE Vaccine Effectiveness Expert Panel subgroup [9].

#### 9.4 Comparison between vaccine efficacy assumptions.

We now compare the aggregate results for the three vaccine efficacy assumptions and four models of alternative vaccination priorities (Table S5, Fig. S10). Although there are large and profound differences between the assumptions for vaccine efficacy, by refitting each to match the observed epidemic we considerably reduce the variability in our results.

**Table S5:** Deaths and hospital admissions in England from 8th December 2020 to 1st September 2021, as observed (top row) and from four model scenarios and four sets of vaccine efficacy parameters (default as in the main paper, and mid-point estimate, low and high confidence intervals as determined by [9]. We give the mean and 95% prediction intervals for each measure. All model results are from 400 samples of the posterior distribution.

| Model | VE | Deaths | Hospital Admissions |
|---|---|---|---|
| Observed | | 62163 | 243573 |
| (i) Default | default | 61,200 (59,900-62,400) | 245,800 (243,500-247,900) |
| | mid | 61,100 (60,200-62,000) | 242,600 (240,000-244,600) |
| | low | 61,300 (59,200-62,600) | 243,000 (239,000-247,200) |
| | high | 61,100 (58,900-63,000) | 238,000 (230,600-242,400) |
| (ii) Prioritise youngest | default | 84,500 (71,200-163,600) | 261,400 (223,600-487,100) |
| | mid | 82,400 (71,900-140,200) | 256,600 (223,900-380,900) |
| | low | 81,400 (71,500-122,700) | 259,300 (226,800-375,600) |
| | high | 81,600 (70,300-155,200) | 245,900 (215,500-404,900) |
| (iii) 3-week interval, default efficacy | default | 69,300 (67,000-71,100) | 314,100 (302,800-329,300) |
| | mid | 69,000 (67,300-71,100) | 304,800 (294,100-319,900) |
| | low | 68,900 (66,400-71,300) | 293,800 (283,500-303,800) |
| | high | 70,600 (66,900-74,400) | 313,800 (297,300-334,700) |
| (iv) 3-week interval, lower efficacy | default | 74,900 (70,700-79,900) | 341,400 (322,400-368,600) |
| | mid | 72,100 (69,800-75,300) | 324,500 (310,100-344,700) |
| | low | 72,100 (68,700-75,700) | 314,300 (300,000-327,900) |
| | high | 74,700 (70,200-79,700) | 336,900 (315,900-363,500) |

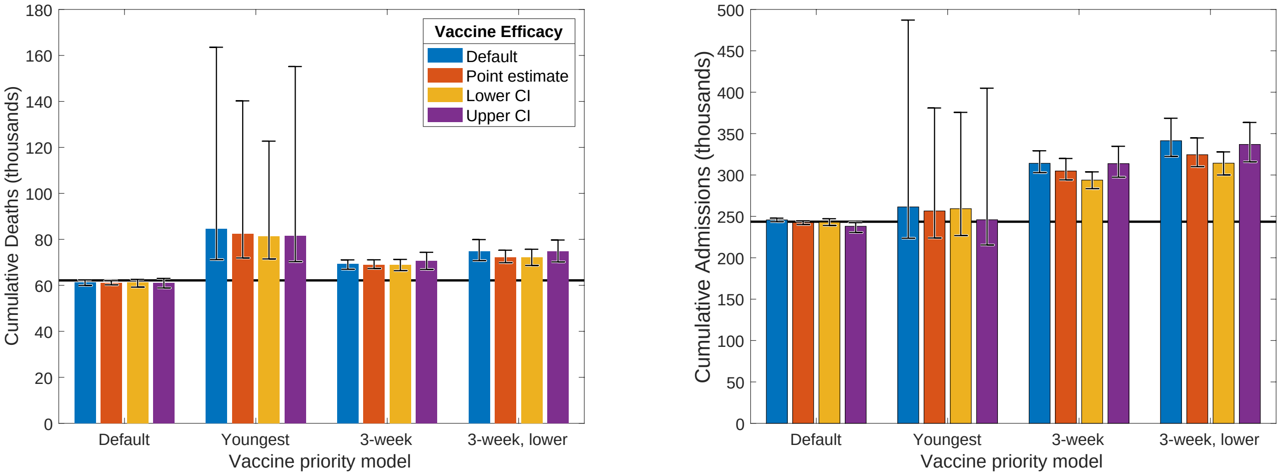


**Fig. S10:** Deaths and hospital admissions in England from 8th December 2020 to 1st September 2021. The four models for vaccine priority are clustered on the x-axis, while colours represent the four vaccine efficacy papers; the horizontal line shows the observed value. Bar heights correspond to the mean and the whiskers to the 95% prediction interval (spanning the 2.5th to 97.5th percentiles) for each measure. These results reflect the values in Table S5.
